# Antidopaminergic Medications Are Associated with Faster Decline in Measures of Clinical Outcome in HD: Insights from PROOF-HD

**DOI:** 10.1101/2025.10.30.25339054

**Authors:** Karen Elta Anderson, Andrew M. Tan, Andrew Feigin, Ralf Reilmann, Anne E. Rosser, Lynn A. Raymond, Sandra K. Kostyk, Carsten Saft, Kelly Chen, Randal Hand, Michal Geva, Michael R. Hayden

## Abstract

**Background:** Antidopaminergic medications (ADMs), including vesicular monoamine transporter-2 (VMAT2) inhibitors and antipsychotics, are frequently-used to manage Huntington disease (HD) symptoms. Prior studies suggest that ADMs may be associated with worsening on measures of outcome in HD clinical trials. The PROOF-HD placebo arm (NCT04556656) provided a controlled, double-blind setting to evaluate ADM impacts on measures of HD progression.

**Objective:** Assess the association between ADM exposure and change in clinical outcomes in the placebo arm of PROOF-HD.

**Methods:** Placebo-arm participants (n=247) were categorized as on- vs off-ADMs. Overall main analyses were corroborated by propensity-score weighting (PSW)-adjusted analyses. Unadjusted analyses examined exposure by ADM class and dose. Outcomes included Total Functional Capacity (TFC), composite Unified Huntington’s Disease Rating Scale (cUHDRS), Stroop Word Reading (SWR), Symbol Digit Modalities Test (SDMT), and Total Motor Score (TMS).

**Results:** Group differences (Δ) favored off-ADMs in cUHDRS (Weeks 39–78) and TFC (Weeks 26–78); at Week 52, cUHDRS had Δ=0.66 (95% CI 0.31–1.01; p=0.0002) and TFC with Δ=0.85 (95% CI 0.47–1.22; p<0.0001) as compared with on-ADMs. Other outcomes were significant or directionally-favored off-ADM participants beyond Week 39. All TMS-subdomain scores, except for chorea, directionally-favored off-ADMs at all visits. Antipsychotic-only and higher-dose ADMs were associated with worse cUHDRS and TFC vs off-ADMs.

**Conclusions:** In this *post hoc* study, ADM use was associated with greater worsening of measures of global, functional, cognitive, and motor outcomes, versus off-ADMs. Accounting for ADM exposure and dose is essential for the interpretation of results from HD trials.

## Introduction

Huntington’s disease (HD) is a progressive neurodegenerative disease caused by a CAG repeat expansion in the HTT gene, leading to widespread neuronal dysfunction and a predictable 10–20 year decline in functional independence that culminates in premature mortality [1–3]. Currently, only symptomatic treatments are available, including vesicular monoamine transporter-2 inhibitors (VMAT2i; i.e., tetrabenazine, deutetrabenazine, and valbenazine), approved for chorea in HD, and off-label use of antipsychotics (e.g., olanzapine, risperidone, aripiprazole, quetiapine) frequently used in clinical practice for managing chorea and psychiatric symptoms, together called antidopaminergic medications (ADMs) [4–11]. Other off-label agents include antidepressants and anxiolytics [12–14]. Emerging evidence suggest that ADM use in particular may be associated with worsening measures of outcomes used in HD clinical trials.

Clinically relevant measures such as Total Functional Capacity (TFC) [15, 16] and the composite Unified Huntington’s Disease Rating Scale (cUHDRS) [17], which integrates motor, cognitive, and functional outcomes into a single global score, overlap with the side-effect profiles of ADMs, potentially mimicking or masking disease symptoms in trials. Some documents suggest that, in practice, ADMs may improve chorea yet worsen non-chorea Total Motor Score (TMS) subdomains (hand and eye movements, dysarthria, bradykinesia) [18, 19]. Dose-dependent risks for cognitive impairment, sedation, and extrapyramidal symptoms (EPS) including parkinsonism and bradykinesia may further affect clinical assessments. Pharmacokinetic factors (e.g., CYP2D6 metabolism) and the strong D2 antagonism associated with many ADMs also complicate clinical care and interpretation of trial assessments [20–24].

Observational data from longitudinal cohorts (ENROLL-HD, REGISTRY) support this concern, showing faster functional and cognitive decline with ADM exposure (e.g., TFC, SWR) [18, 19, 25]. A 12-week placebo-controlled randomized trial demonstrated that tetrabenazine improved chorea but worsened function and cognition versus placebo [26]. Yet, to date, no randomized controlled trial has prospectively tested whether ADM use alters outcome trajectories or biases measures of clinical outcome.

The Phase 3 PROOF-HD placebo arm provided a well-controlled framework to explore the impact of ADM use on key clinical measures of HD progression. Building on published findings [18, 19, 21, 25], we performed a *post hoc* analysis to assess whether ADM exposure was associated with different longitudinal outcome scores across TFC, cUHDRS, SDMT, SWR, and TMS (including subdomains). Our main findings have implications for both HD trial design and therapeutic interventions in populations where ADM use is common.

## Methods

### Study Design, Participants, and Population Definitions

This *post hoc* analysis was conducted using data from the placebo arm of the Phase III PROOF-HD trial (NCT04556656; EudraCT 2020-002822-10), a multicenter, randomized, double-blind, placebo-controlled study designed to evaluate the efficacy and safety of pridopidine in patients with early-stage HD. Eligibility criteria were defined in the PROOF-HD protocol [27] and are available in the published protocol and on ClinicalTrials.gov (NCT04556656; accessed October 6, 2025). The study enrolled 499 participants across 59 international sites [27]. Within the analyzed placebo arm (n=247, mITT population), 135 (54.7%) were on ADMs anytime during the study and 112 (45.3%) were off ADMs.

For analyses, participants were categorized based on ADM use during the study:

- **On-ADMs:** Participants who received a VMAT2 inhibitor and/or antipsychotics at any time during the double-blind treatment period, regardless of whether use began at baseline or post-baseline.
- **Off-ADMs:** Participants who were not prescribed VMAT2 inhibitors or antipsychotics at any time during the double-blind treatment period, including at baseline and throughout follow-up.

Dose thresholds, higher and lower, for each ADM were defined based on regulatory (FDA) labeling (half the daily dose) and established pharmacodynamic considerations (**Supplemental Table 1**). For all ADMs, “lower dose” followed recommended dose-reduction guidance when co-administered with strong CYP2D6 inhibitors. For antipsychotics, “lower dose” also corresponded to exposures below levels associated with ∼75–80% D2 receptor occupancy, above which extrapyramidal symptoms and cognitive impairment increase [4, 9, 23, 28]. Doses above these cutoffs were classified as “higher dose.”

### Clinical Outcome Measures

Clinical outcomes were evaluated to assess the impact of ADM use in participants randomized to the placebo arm of the PROOF-HD trial. We assessed HD outcome measures, including TFC, SDMT, SWR, TMS, and the cUHDRS. TFC tracks loss of daily function [3, 15, 29]. SDMT and SWR index processing speed/executive function and attention; TMS captures motor signs and symptoms; cUHDRS integrates these outcomes, and was selected based on its sensitivity and superior ability to detect clinical decline in early HD vs each of its individual components (TFC, SDMT, SWR, TMS) [17, 30]. We additionally evaluated VMAT2i-only and antipsychotic-only exposure and dose effects [31–36].

### Statistical Analysis

All statistical analyses were performed using SAS version 9.4 or higher [27]. The primary analysis utilized a mixed-effects model for repeated measures (MMRM) to assess longitudinal changes in clinical outcomes between participants off-ADMs versus those on-ADMs in the mITT populations. The MMRM approach was selected to account for within-subject correlations, and to handle missing data under the assumption that data were missing at random (MAR).

Longitudinal changes from baseline in all endpoints were modeled as a function of ADM use (on vs. off), visit (Weeks 26, 39, 52, 65, and 78), baseline value of endpoint in analysis, treatment (placebo, pridopidine), region (Europe, North America), HD stage as defined by randomization strata, and 3 interaction terms: treatment-by-visit, treatment-by-ADM use, and ADM use-by-visit. In subsequent analyses, ADM use was replaced by antipsychotic- or VMAT2i-use to evaluate their respective association with clinical outcomes using the same modeling approach.

In the placebo arm of PROOF-HD, the dropout rate was ∼8.9% [27]. These dropout patterns were considered when addressing potential biases related to missing data and differential study retention across treatment groups.

To account for the potential effect of baseline differences, a sensitivity analysis using a propensity score weighting (PSW) method with 10 covariates—age, sex, region, CAG repeats, CAP100 (calculated as age x (CAG-30)/6.49), TFC, TMS, SWR, SDMT, and Q-Motor FT-IOI mean—to adjust for baseline differences between the on- and off-ADM groups in key metrics related to HD progression (see **Table 1, Supplemental Materials**, **Supplemental Figure 1** and **Supplemental Figure 2**).

**Table 1.** Baseline characteristics of placebo participants on-versus off-ADMs (mITT). Baseline demographic and clinical measures for placebo-arm participants categorized by ADM exposure. Values are shown unadjusted and after propensity score weighting (PSW) using 10 covariates (age, sex, region, CAG repeat length, CAP100, TFC, TMS, SWR, SDMT, Q-Motor FT-IOI mean). On-ADM is ADM use anytime during the study. Off ADMs is unexposed all the time during the study. *Two patients on-ADMs were trimmed as their PS scores were below 5th percentile.

|  |  | Unadjusted |  | Propensity Score Weighted |  |
| --- | --- | --- | --- | --- | --- |
|  | Total<br>N=247 | Off ADM<br>N=112 | On ADM<br>N=135 | Off ADM<br>N=112 | *On ADM<br>N=133 |
| **Duration since Symptoms (y) Onset, Mean (SD)** | 4.6 (4.6) | 4.1 (4) | 5.1 (5) |  |  |
| **HD1 (TFC 11-13), n(%)** | 102 (41.3%) | 57 (50.9%) | 45 (33.8%) |  |  |
| **HD2 (TFC 7-10), n(%)** | 145 (58.7%) | 55 (49.1%) | 88 (66.2%) |  |  |
| **Age (y), mean (SD)** | 52.6 (11.4) | 52.3 (11.3) | 53.1 (11.5) | 52.3 (11.3) | 52.9 (10.5) |
| **Female n (%)** | 126 (51%) | 62 (55.4%) | 64 (48.1%) | 62 (55.4%) | 69 (51.9%) |
| **CAG, mean (SD)** | 43.6 (3.3) | 43.1 (3) | 44.1 (3.5) | 43.1 (3) | 43.2 (3.1) |
| **CAP (100), mean (SD)** | 105.8 (15.4) | 101.5 (15.3) | 109.7 (14.5) | 101.5 (15.3) | 103.4 (15.3) |
| **TFC, mean (SD)** | 9.9 (1.7) | 10.3 (1.6) | 9.5 (1.7) | 10.3 (1.6) | 10.3 (1.6) |
| **cUHDRS, mean (SD)** | 8.9 (2.7) | 9.9 (2.4) | 8.0 (2.5) | 9.7 (2.2) | 9.7 (2.2) |
| **TMS, mean (SD)** | 32.9 (10.9) | 30.3 (9.6) | 34.9 (11.6) | 30.3 (9.6) | 31.1 (9.9) |
| **SDMT, mean (SD)** | 23.3 (9.4) | 26.8 (9.7) | 20.4 (8.2) | 26.7 (9.4) | 25.5 (7.5) |
| **SWR, mean (SD)** | 61.9 (18.2) | 68.8 (16.6) | 56.3 (17.7) | 68.9 (16.4) | 68.7 (17.7) |

To evaluate dose-dependent effects of VMAT2i on the clinical outcomes, an MMRM similar to the primary model was applied, replacing the binary VMAT2i use variable with a 3-level categorical variable (recommended label dose range, i.e., lower or higher dose, and off-ADMs). Owing to small samples, these models were unadjusted (no PSW). For descriptive interpretability of MMRM estimates, **Supplemental Table 4** summarizes the percent (%) reduction in decline off- vs on-ADMs using model-derived changes. The metric was defined as 100% × (1 − R), where R is the ratio of baseline-adjusted decline between ADM exposure groups. Baseline adjustment was implemented by scaling each group’s LS mean change from baseline by its corresponding group baseline mean. All statistical tests were two-sided, with a significance threshold of α=0.05 and reported with 95% confidence intervals (CIs).

### Ethics

This analysis used de-identified data from the Phase 3 PROOF-HD trial [27], which was conducted in accordance with the Declaration of Helsinki and ICH-GCP guidelines, with IRB/IEC approval at all participating sites and written informed consent from all participants. As this study involved secondary analysis of existing de-identified data, no new ethical approval was required from the ethics committees for the current report.

## Results

### Participant Characteristics and ADM Use in the PROOF-HD Placebo Arm

In PROOF-HD, 54.7% (135/247) of participants in the placebo group received at least one ADM anytime during the study and 112 (45.3%) were off-ADMs all the time during the study (**Supplemental Table 2**). There were 87 (35.2%) subjects on antipsychotics-only and 29 (11.7%) on VMAT2i-only. A small number of patients (7.7%) were using >1 ADMs at the same time (**Supplemental Table 2**). Among antipsychotic users, olanzapine (10.9%), risperidone (10.5%), tiapride (7.3%), aripiprazole (5.3%), and quetiapine (4.9%) were the most common. VMAT2i use was split between deutetrabenazine (6.9%) and tetrabenazine (4.9%) (**Supplemental Table 3**).

At baseline, participants on-ADM showed slightly more advanced disease as compared with off-ADMs (**Table 1**). To assess this imbalance, propensity score weighting (PSW) was applied using 10 covariates (**Methods**). After PSW, baseline balance was achieved (e.g., TFC 10.3 both; cUHDRS 9.7 both; SDMT 26.7 vs 25.5; SWR 68.9 vs 68.7; TMS 30.3 vs 31.1), supporting interpretable between-group comparisons.

### Impact of ADM Use on Worsening Measures of Clinical Outcome

To evaluate whether ADM exposure influenced the longitudinal trajectory of clinical decline, we assessed change from baseline across outcome measures in the placebo arm of the PROOF-HD trial (on-ADM vs off-ADM) (**Figure 1**). Complete numerical results are provided in the **Supplemental Materials.**

**Figure 1.**
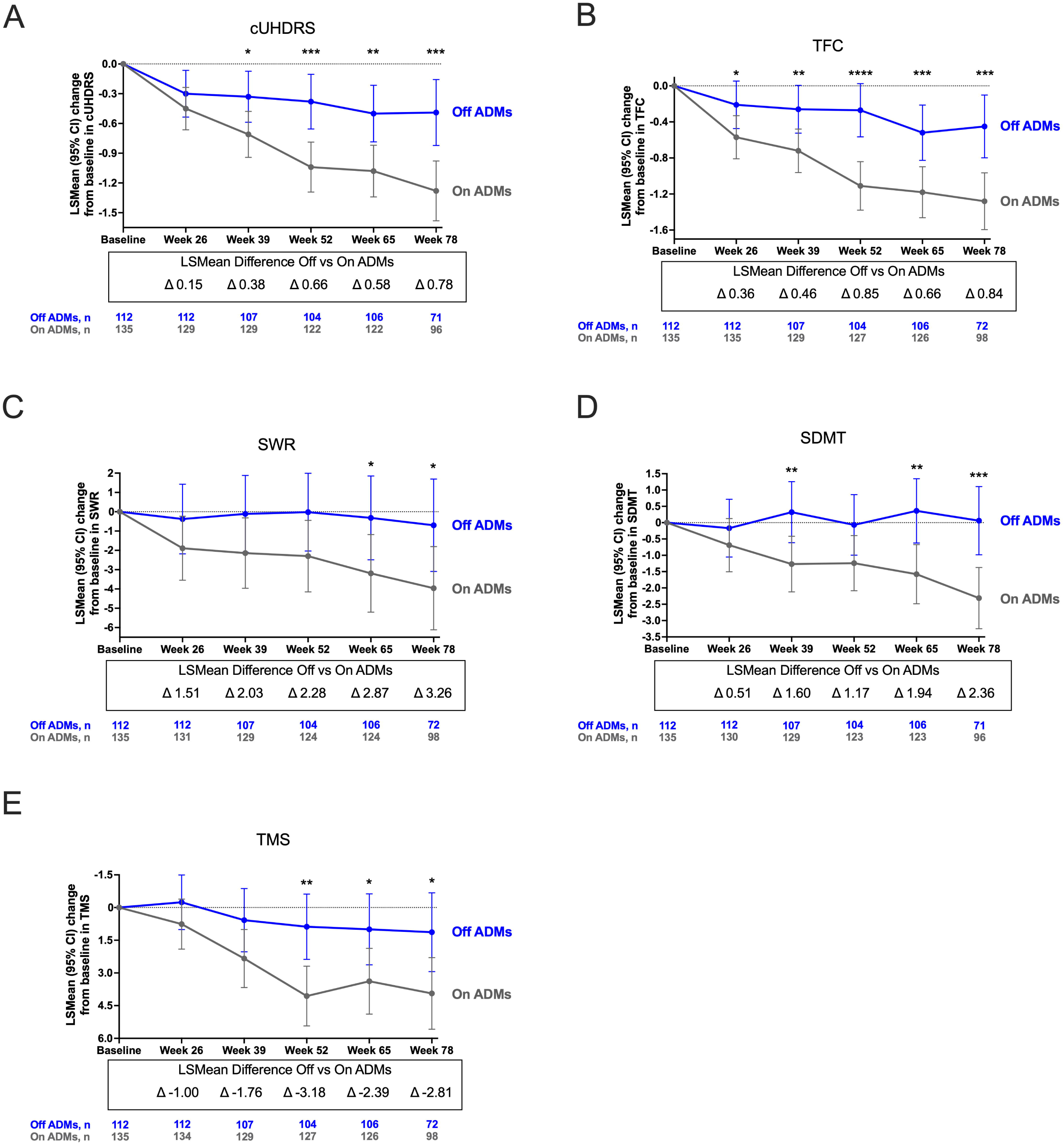
Longitudinal changes in clinical outcomes by ADM exposure. LS mean change (Δ) from baseline with 95% CI for (A) cUHDRS, (B) TFC, (C) SWR, (D) SDMT, and (E) TMS in placebo participants on-ADMs vs off-ADMs. Positive Δ favors off-ADM except for TMS, where negative Δ favors off-ADM. * p<0.05; ** p<0.01; *** p<0.001; **** p<0.0001.

In cUHDRS (**Figure 1A**), the on-ADM group demonstrated a faster decline as compared with off-ADMs. Between-group differences emerged by Week 39 and persisted through study end. Differences became significant at Week 39 (p=0.03), Week 52 (p=0.0002), Week 65 (p=0.002), and Week 78 (p=0.0002). On-ADM decline worsened vs off-ADMs by 62.6% at Week 65 and 69.1% at Week 78.

In TFC (**Figure 1B)**, decline was consistently worse on-ADMs. The difference between groups was significant from Week 26 onward (p=0.04), increasing at Week 39 (p=0.01), peaking at Week 52 (p<0.0001), and remaining significant through Week 65 (p=0.0008) and Week 78 (p=0.0001). On-ADM participants showed faster functional decline across all timepoint with a decline of 59.4% at Week 65 and 67.6% at Week 78.

For SWR (**Figure 1C**), group separation trended yet non-significant at Weeks 26 through Week 52; and reached significance at Week 65 (p=0.04) and Week 78 (p=0.03). Off-ADM use corresponded to less SWR decline: 91.8% at Week 65 and 85.6% at Week 78. In SDMT (**Figure 1D**), the off-ADM group had significantly less decline at Week 39 (p=0.0098), which strengthened at Week 65 (p=0.0026) and Week 78 (p=0.0004) vs on-ADMs. Off-ADMs presented with greater decline by 117.4% at Week 65 and 102.0% at Week 78.

For TMS (**Figure 1E**), on-ADM participants showed faster worsening of measures of motor function at later time points with significance appearing at Week 52 (p=0.0010), Week 65 (p=0.02), and Week 78 (p=0.01). On-ADM showed greater TMS worsening by 65.8% at Week 65 and 66.9% at Week 78.

To address the possible effect of baseline imbalance on these observations, we conducted a PSW analysis (see baseline data shown in **Table 1**). Results were directionally consistent with the unadjusted findings, with mid-to-late separation remaining significant for cUHDRS and TFC. Full estimates are shown in **Supplemental Figure 1** and **Supplemental Materials**. Forest plots (**Supplemental Figure 2**) further show the magnitude and directionality of PSW-adjusted between-group differences, consistently favoring off-ADM over on-ADM across all visits and measured outcomes.

#### Effect of ADM Exposure on TMS Subdomains

Many ADMs are prescribed for the management of motor disturbances, particularly chorea. Here, we evaluated ADM exposure across TMS-subdomains (**Figure 2**). In the forest plot, effects favored off-ADMs for all sub-scores, up to Week 52. Hand movements, eye movements, bradykinesia, and dysarthria show the clearest, most durable shifts favoring off-ADMs at Weeks 52, 65, and 78. Interestingly, in contrast, chorea scores favored on-ADMs at Week 65 and 78, though nonsignificant (p>0.05). Class-specific impacts of VMAT2-only (**Supplemental Figure 3**) or antipsychotic-only use (**Supplemental Figure 4**) also show a nonsignificant tendency toward improved chorea symptoms as compared with off-ADMs at later time points, e.g., Week 65 and 78. Both ADM classes showed broader, worse non-chorea motor scores as compared with the off-ADM cohort.

**Figure 2.**
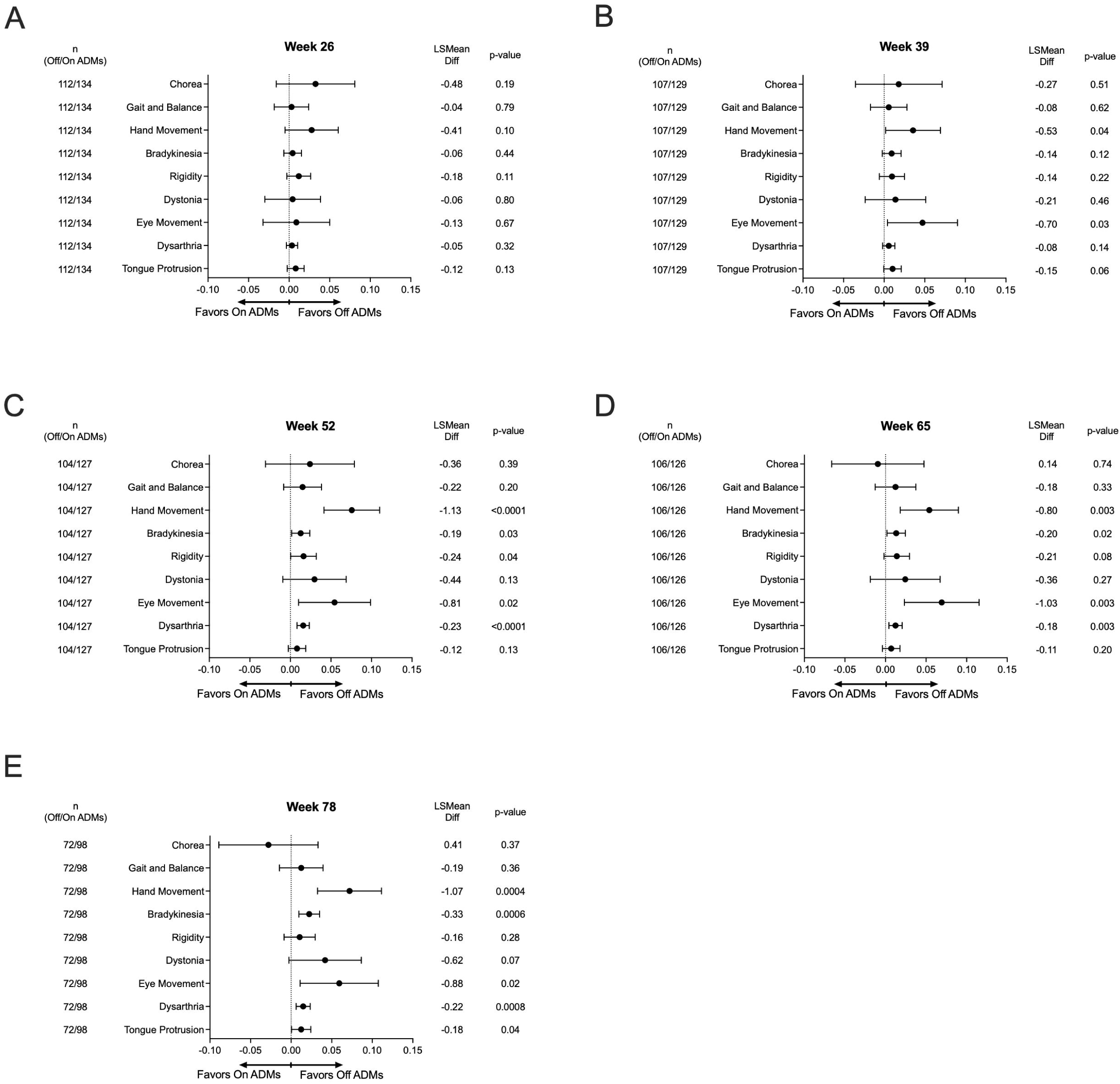
Effect of ADM exposure on TMS subdomains. LS mean differences (Δ off-ADM minus on-ADM) with 95% CI across TMS subdomains. Negative Δ indicates less motor worsening off-ADMs. Class-specific subdomain analyses are shown in **Supplemental Figures 3 and 4.**

### Impact of ADM Class on Measures of Clinical Outcome: VMAT2 inhibitors and Antipsychotics

#### VMAT2 inhibitor Impact

As shown in **Figure 3**, analysis of ADM class-specific exposure were pairwise-compared with off-ADM participants. In cUHDRS (**Figure 3A**), separation favored off-ADMs as compared with VMAT2i-use, and reached significance at Week 52 (p=0.044). Other visits showed a similar, non-significant trend.

**Figure 3.**
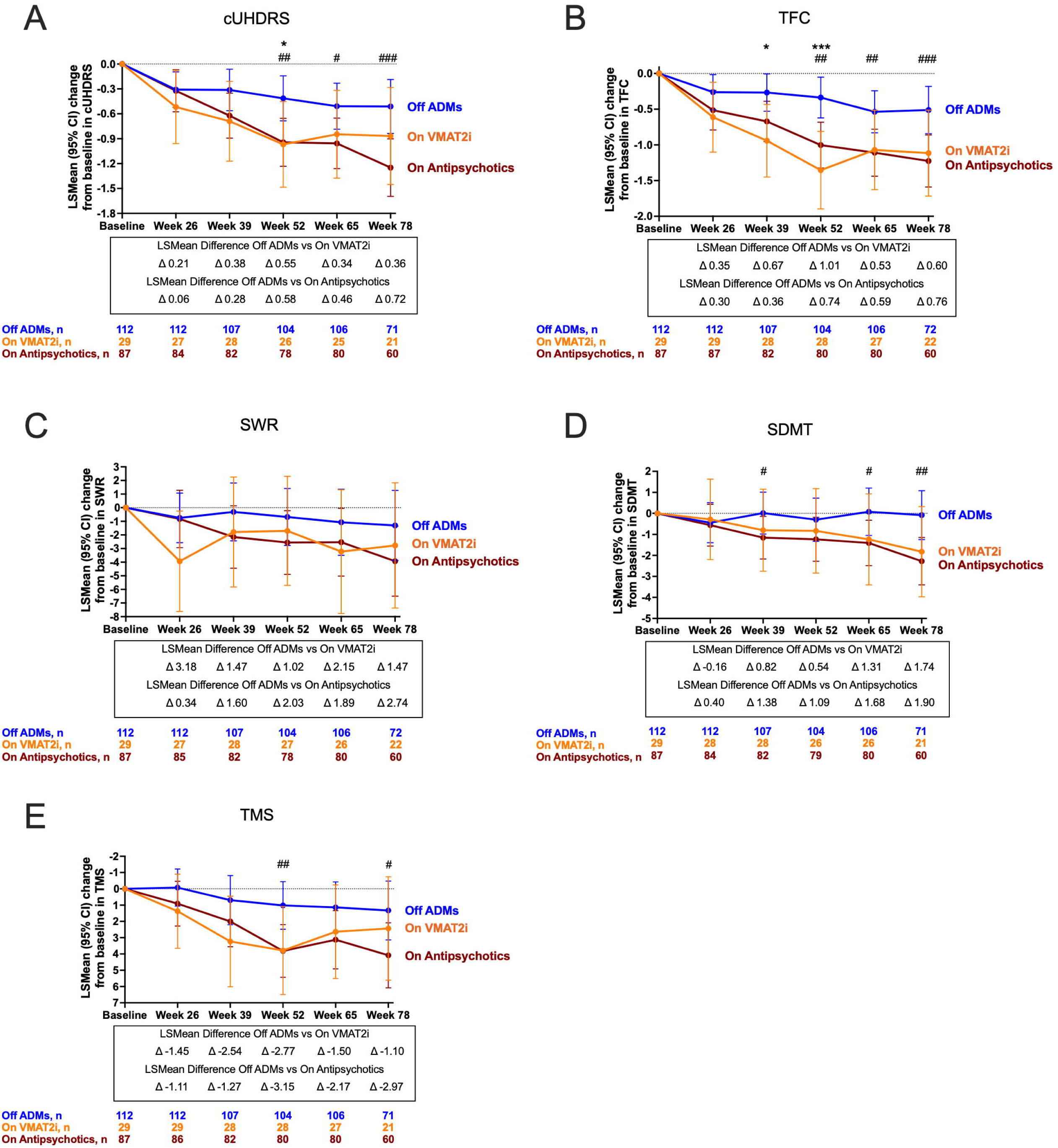
Class-specific ADM exposure and clinical progression. Unadjusted LS mean change from baseline with 95% CI for (A) cUHDRS, (B) TFC, (C) SWR, (D) SDMT, and (E) TMS comparing VMAT2 inhibitor–only, antipsychotic-only, and off-ADM participants. **Supplemental Figures 3 and 4** present class-specific forest plots. Statistical annotations denote pairwise contrasts vs off ADM: asterisks (*) for off ADM vs VMAT2 inhibitor–only and hash symbols (#) for off ADM vs antipsychotic-only. Significance thresholds: * p<0.05; ** p<0.01; *** p<0.001; **** p<0.0001; # p<0.05; ## p<0.01; ### p<0.001; #### p<0.0001.

TFC showed a similar pattern in comparisons of VMAT2i vs off-ADMs (**Figure 3B**). Significant differences favored off-ADMs at Week 39 (p=0.015) and Week 52 (p=0.00057). Earlier and later visits were directionally consistent without significance. Cognitive outcomes, SWR and SDMT (**Figure 3C–D**) were directionally aligned, with less decline off-ADMs as compared with VMAT2i use across visits. Separation did not reach significance at any timepoint, though later visits trended further in the off-ADM direction, likely due to small sample size on VMAT2i only group. TMS motor function (**Figure 3E**) also favored the off-ADM group across visits but was nonsignificant at any visit. Forest plots show a consistent favoring off-ADMs vs VMAT2i for all measures and visits (**Supplemental Figure 5)**, with the largest separations between Weeks 39– 52.

#### Antipsychotics Impact

In cUHDRS (**Figure 3A**), antipsychotic-only exposure resulted in greater decline as compared with off-ADM. Differences emerged at Week 39, and later strengthened, significantly favoring off-ADM at Week 52 (p=0.0018), Week 65 (p=0.017) and Week 78 (p=0.00091). TFC showed the same pattern (**Figure 3B**) with faster decline on antipsychotics versus off-ADMs. Effects were significant at Week 52 (p=0.0003), Week 65 (p=0.0049), and Week 78 (p=0.0008).

Antipsychotic exposure was associated with greater cognitive decline on SDMT, with directionally similar but non-significant differences on SWR (**Figure 3C–D**; see **Supplemental Materials**). In SDMT, significant separation favored off-ADM at Week 39 (p=0.037), Week 65 (p=0.015), and Week 78 (p=0.008). Finally, motor decline was faster on antipsychotics at later time points, with higher (worse) TMS scores versus off-ADMs **(Figure 3E)**. Differences were significant at Week 52 (p=0.0022) and Week 78 (p=0.015), with a borderline effect at Week 65 (p=0.052).

Antipsychotic-only participants showed directional effects that favored off-ADMs across measures in forest plots (**Supplemental Figure 6**), with the most pronounced separations later in the trial post-Week 26 which persisted to study end. Taken together, both VMAT2 inhibitors- and antipsychotic-only exposure were associated with greater decline in global and functional outcomes as compared with off-ADMs. Note that direct class-to-class comparisons were not performed, due in part to smaller on-ADM cohorts which limited power and thus warrant cautious interpretation of these findings.

#### Dose-Dependent Effects of ADMs on Clinical Outcomes

ADM effects are dose-dependent [9, 18, 24, 37, 38]. Participants on higher-dose or lower-dose ADMs (ie, tetrabenazine, deutetrabenazine, quetiapine, aripiprazole, olanzapine, risperidone; thresholds shown in **Supplemental Table 1**) were compared with off-ADMs (**Figure 4**). In cUHDRS (**Figure 4A**), higher-dose ADMs significantly declined as compared with off-ADM beginning at Week 39 (p=0.0449) and remained significant through Week 52 (p=0.0003), Week 65 (p=0.0012), and Week 78 (p=<0.0001). Lower-dose ADMs was not-significant compared with off-ADMs across visits. TFC measures followed a similar pattern with higher-dose ADMs showing significant differences as compared with off-ADM at all visits; at Week 26 (p=0.0024), Week 39 (p=0.0007), Week 52 (p<0.0001), Week 65 (p<0.0001), and Week 78 (p<0.0001) (**Figure 4B**). Lower-dose ADMs effect sizes remained small and non-significant across visits as compared with off-ADMs.

**Figure 4.**
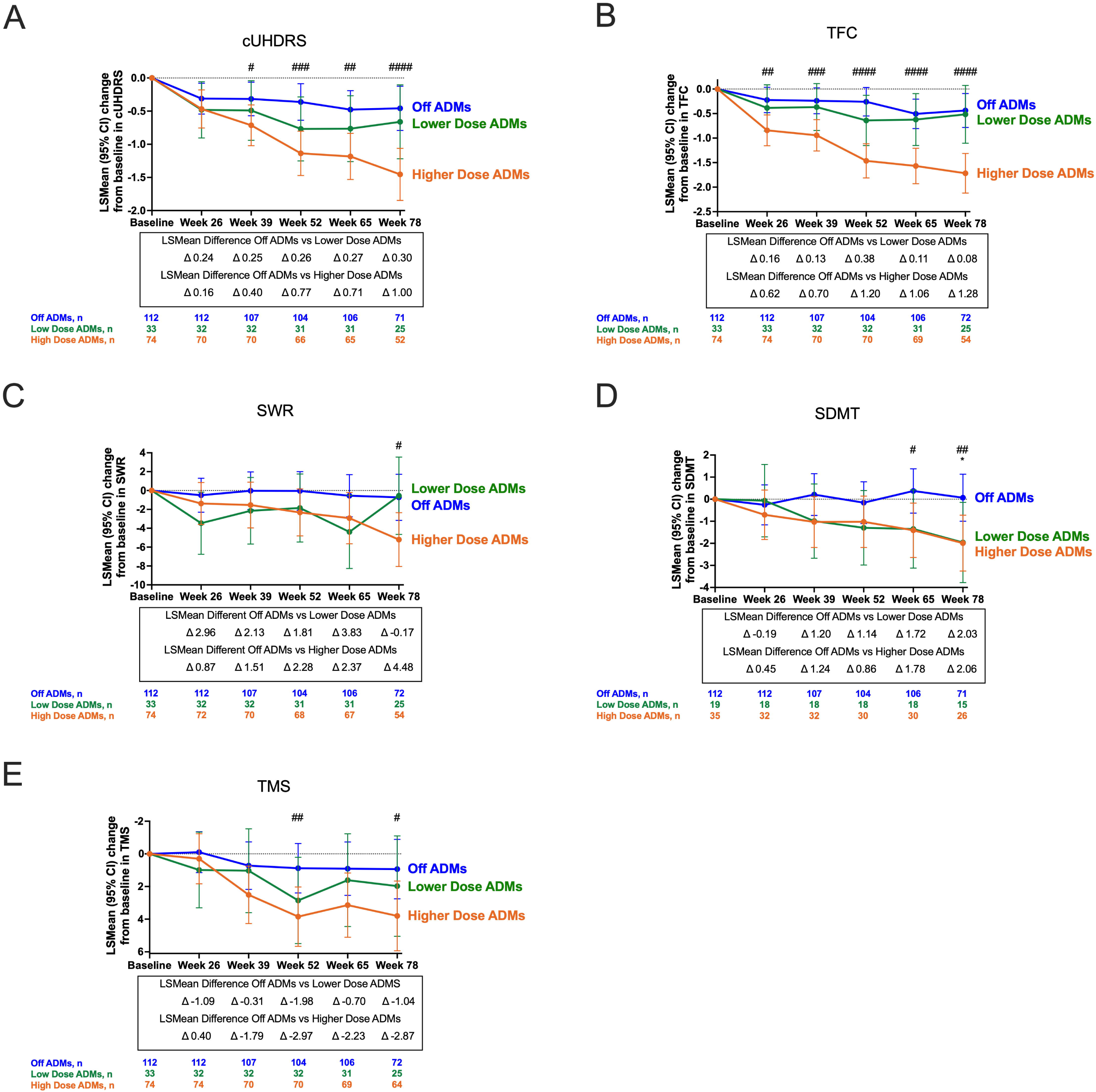
Dose-dependent effects of ADM exposure on clinical outcomes. Unadjusted LS mean differences (Δ off-ADM minus on-ADM group) with 95% CI for (A) cUHDRS, (B) TFC, (C) SWR, (D) SDMT, and (E) TMS comparing higher-dose ADMs, lower-dose ADMs, and off-ADM. Dose thresholds are provided in **Supplemental Table 1**. Statistical annotations denote pairwise contrasts vs off ADM: asterisks (*) for off ADM vs lower-dose ADMs and hash symbols (#) for off ADM vs higher-dose ADMs. Significance thresholds: * p<0.05; ** p<0.01; *** p<0.001; **** p<0.0001; # p<0.05; ## p<0.01; ### p<0.001; #### p<0.0001.

Both cognitive measures, SWR and SDMT, reached significance only later in the study for higher-dose ADMs as compared with off-ADM. For SWR, higher-dose ADMs became significant at Week 78 (p=0.0106), showing a worsening trend from early visits to the end of the trial (**Figure 4C**). Similarly, lower-dose ADMs showed variable, non-significant differences as compared with off-ADMs. As with SWR, the higher-dose ADM group in the SDMT measure became significant at Weeks 65 (p=0.0214) and 78 (p=0.0094), while lower-dose ADMs reached significance at Week 78 (p=0.0413) (**Figure 4D**).

For TMS, where negative Δ indicates less motor worsening off-ADMs (**Figure 4E**), higher-doses showed greater worsening versus off-ADM, with significance at Weeks 52 (p=0.0087) and Week 78 (p=0.0264). TMS scores for other visits were inconsistent and non-significant. Lower-dose ADMs showed no significant separation from off-ADMs at any visit.

Overall, higher-dose ADM exposure was associated with greater worsening over time as compared with off-ADM. In contrast, lower-dose ADM exposure tracked closely with off-ADM across outcomes.

## Discussion

The impact of ADMs on the natural progression of HD is not well-understood, despite their longstanding and widespread use in clinical practice. Placebo-controlled trials have not prospectively quantified ADM exposure and its association with longitudinal outcomes under randomized trial monitoring. Emerging evidence suggests that ADMs may be associated with worsening measures of clinical outcome in HD [19–21, 24, 25, 39].

In the present report, we investigated how ADM use within the PROOF-HD placebo group influenced clinical outcomes over 78 weeks. Among participants, ADM use was associated with a greater decline in functional, cognitive, and motor outcomes, with significant differences emerging at Week 26 for TFC, at Week 39 for cUHDRS and SDMT, and at Week 52 and Week 65 for TMS and SWR, respectively, with effects persisting to Week 78. Propensity score weighting of these comparisons confirmed these patterns.

Across dose analyses, higher-dose ADMs were associated with greater worsening on cUHDRS and TFC. For cognition, both doses performed similarly, and for TMS the higher dose was worse than lower dose; both worse than off-ADMs. VMAT2i-only dose findings mirrored this dose– response for cUHDRS and TFC; cognitive effects were directionally negative but non-significant, and TMS signals were inconsistent. Antipsychotic-only dose cohorts were too small for stable estimates, but a prior causal analysis suggests dose-dependent associations with greater functional and cognitive worsening at higher doses as compared with lower dose and off-ADM [18]. Collectively, these results are compatible with a dose–response relationship and with prior pharmacodynamic work indicating that small dose increases—particularly with high-affinity D2 antagonists, e.g., risperidone—can produce disproportionately large clinical effects [9, 28, 40]. Together, our study provides clinical evidence that extends prior observations showing that ADM use may worsen measures of outcome in HD.

Our findings should be interpreted with caution given several caveats. Subgroup analyses were power-limited and focused on commonly used ADMs. To simplify interpretation, participants simultaneously receiving an antipsychotic and a VMAT2i were excluded from class-specific analyses. In this regard, prior reports show that multi-ADM use is common and associated with greater worsening of clinical measures; thus, concurrent use would be expected to bias estimates toward greater decline [20, 24, 41–43].

ADMs are often initiated for behavioral symptoms (e.g., agitation, psychosis, irritability) that can reflect more advanced disease. As such, confounding by intention/indication, where ADMs are started or escalated for worsening symptoms, may contribute to the observed associations. For example, Zeng et al. (2025) reported that individuals at clinical high risk for psychosis who received atypical antipsychotics had more severe baseline symptoms and poorer function than unmedicated individuals [10]. Our analyses addressed baseline imbalance using multiple disease-severity indicators and PSW. Due to limited power, PSW was not used for ADM-class or dose-specific analyses. Nevertheless, effect directions for TFC and cUHDRS were consistent with the unadjusted and baseline-adjusted findings, supporting an association between ADM exposure and faster decline as compared with off-ADM.

In agreement with our present report, retrospective, observational, and causal modeling studies independently suggest that ADM exposure is associated with worsened measures of clinical outcome [24, 39]. In REGISTRY, ADM-treated participants declined faster in TFC and other functional measures, a pattern not fully explained by baseline severity [20]. Matched analyses from Enroll-HD showed that ADM exposure alleviated some behavioral symptoms but was associated with worsening of functional and cognitive measures as compared with off-ADM, with accelerated cognitive decline observed shortly after ADM initiation [21].

Predictive and causal modeling studies further support plausibility. In Ghazaleh et al. [25], ADM use was among the top predictors of cognitive and functional decline across >1,600 Enroll-HD participants after adjustment for >100 baseline variables. Boareto et al. [19] likewise identified baseline ADM use as one of the strongest independent predictors of progression across TFC, TMS, SDMT, and SWR. A causal-inference analysis in Enroll-HD, adjusting for 27 covariates, reported a dose-dependent association between ADM exposure and worsening over two years, arguing against baseline imbalance as a sole explanation [18]. Collectively, these convergent data implicate ADM exposure in poorer measures of functional and cognitive decline in HD.

In routine practice, distinguishing medication effects from disease progression is challenging. This is reflected in the FDA label for both tetrabenazine and AUSTEDO (deutetrabenazine); where it states that “…*it may be difficult to distinguish between adverse reactions and progression of the underlying disease; decreasing the dose or stopping the drug may help*” [33].

Our findings strongly suggest that ADM use has the potential to meaningfully influence clinical outcomes in HD trials [27], distorting observed trajectories and obscuring true treatment effects of the trial intervention [18, 20, 39, 44]. Interestingly, this concern may also extend beyond HD. In large schizophrenia cohorts, Dickerson et al. (2025) and Lock et al. (2025) reported associations between antidopaminergic agents (e.g., clozapine, haloperidol, quetiapine) and poorer cognitive performance, particularly in memory, while no comparable associations were observed for antidepressants, consistent with Geva et al., 2025, suggesting that the observed burden is specific to ADMs [18, 45, 46].

Our study is not intended to inform changes to the current standard-of-care or guide clinical decision-making in HD. ADMs remain an important component of symptom management. They also should not be interpreted as a suggestion that patients on ADMs should never be enrolled in clinical trials. Nonetheless, the observed associations support a biologically plausible relationship between ADM exposure and worsening measures of HD outcomes. These results provide essential groundwork for designing and interpreting future trials. Although power-limited, the absence of detectable worsening with lower-dose exposure in several outcomes suggests a potential combinatorial strategy with investigational therapies, balancing symptom control against confounding risk, but this requires confirmatory trial validation. Ultimately, dedicated prospective studies are needed to confirm these associations and to inform trial designs that explicitly account for ADM exposure.

The data presented here do not show that ADMs cause worsening in clinical measures; rather, they demonstrate a consistent association. ADM use may serve as a marker of participants with intrinsically faster progression. Either way, ADM use should be accounted for—preferably by balancing at randomization—and, at minimum, incorporated into analyses and interpretation plans. Imbalances in ADM exposure between active and placebo groups can bias outcomes, yielding false-positive or false-negative conclusions. Our findings strongly suggest that ADM use has the potential to meaningfully influence clinical outcomes in HD trials, distorting observed trajectories and confounding true treatment effects [18, 20, 39, 44]. These factors should be taken into account when designing future studies.

## Supporting information

Dose thresholds for VMAT2 inhibitors and antipsychotics used for dose response subgrouping

ADM exposure frequencies in the placebo arm (mITT).

Antipsychotic and VMAT2i use by agent (non mutually exclusive).

Percent (%) decline off vs on-ADMs (unadjusted and PSW).

Propensity score weighted clinical outcomes by ADM exposure.

PSW adjusted forest plots of visit-wise differences (off-ADM vs on-ADM) across clinical outcomes (95% CI).

TMS subdomains for VMAT2 inhibitor only vs off-ADM (95% CI) forest

TMS subdomains with antipsychotic only vs off-ADM (95% CI) forest plots.

VMAT2 inhibitor only vs off-ADM across outcomes (95% CI) forest plots.

Antipsychotic only vs off ADM across outcomes (95% CI) forest plots.

Supplemental Material (results extended text)

## Supplementary Material (Legends)

**Supplemental Table 1. Dose thresholds for VMAT2 inhibitors and antipsychotics used for dose–response subgrouping.** ADMs are grouped by class: “VMAT2 inhibitors (not on antipsychotics)” includes participants on dTBZ or TBZ only; “Antipsychotics (not on VMAT2i)” includes participants on a single antipsychotic only. Columns list the prespecified lower-dose and higher-dose cutoffs in mg/day with the number of participants in each bin (n). For lower dose, a participant could be on only one ADM at or below the stated cutoff for that agent/class. For higher dose, a participant could be on more than one ADM; assignment to the higher-dose bin was based on any qualifying ADM above the stated cutoff for that agent/class. Counts are mutually exclusive within each class block. These thresholds are the basis for the reported dose-dependent subgroup analyses (shown in **Figure 4**).

**Supplemental Table 2. ADM exposure frequencies in the placebo arm (mITT).** Frequencies and percentages are shown for the PROOF-HD placebo mITT population [27]. “Off ADMs all the time during the study” indicates no exposure to antipsychotics or VMAT2 inhibitors at baseline or at any visit during the double-blind period. “On ADMs anytime during the study” indicates exposure to any ADM at one or more visits. Class-specific rows restrict exposure to a single class only. Class-specific rows exclude the other class (i.e., antipsychotic rows exclude VMAT2i, and VMAT2i rows exclude antipsychotics). Counts represent unique participants. The “on >1 ADM” row is an overlapping subset of “on ADMs anytime” and may include participants on two or more antipsychotics or on a VMAT2i plus an antipsychotic; thus, row percentages need not sum to 100%.

**Supplemental Table 3. Antipsychotic and VMAT2i use by agent (non-mutually exclusive).** Counts and percentages are shown for the total PROOF-HD mITT population (N=247). Values reflect any exposure at any time during the double-blind period. Rows are non-mutually exclusive: A participant can appear in more than one antipsychotic row if they received multiple antipsychotics, and can also appear in both VMAT2 inhibitor rows if they received both agents. Percentages use the total population as the denominator; therefore, row totals do not sum to 100%. “Other” antipsychotics include fluphenazine and haloperidol. VMAT2i, vesicular monoamine transporter-2 inhibitor; mITT, modified intent-to-treat.

**Supplemental Table 4. Percent (%) decline off vs on-ADMs (unadjusted and PSW)**. Values summarize the percent (%) reduction in decline for participants off-ADM versus on-ADM in the PROOF-HD placebo mITT population across visits. Estimates are based on model-derived LS mean change from baseline (see **Methods**). Baseline adjustment was implemented by scaling each group’s LS mean change by its own group baseline mean. To harmonize outcome directions, “decline” was defined as change in the direction of clinical worsening: negative change for cUHDRS, TFC, SWR, and SDMT; positive change for TMS (higher scores = worse). Thus:

- **>0%** indicates less worsening off-ADM than on-ADM.
- **100%** indicates no worsening off-ADM (i.e., off-ADM change ∼0 when on-ADM worsened).
- **>100%** indicates net improvement off-ADM relative to on-ADM worsening.
- **<0%** (if observed) would indicate more worsening off-ADM.

“Unadj” uses the primary (unweighted) model; PSW applies propensity-score weighting to re-weight baseline covariates prior to estimating LS means. Percentages are based on the magnitude of model-estimated changes and may not sum across timepoints; minor differences reflect rounding. ADM, antidopaminergic medication; PSW, propensity-score weighted; mITT, modified intent-to-treat; cUHDRS, composite Unified Huntington’s Disease Rating Scale; TFC, Total Functional Capacity; SWR, Stroop Word Reading; SDMT, Symbol Digit Modalities Test; TMS, Total Motor Score.

**Supplemental Figure 1. Propensity score–weighted clinical outcomes by ADM exposure.** PSW-adjusted LS mean change from baseline (95% CI) for (A) cUHDRS, (B) TFC, (C) SWR, (D) SDMT, and (E) TMS comparing off-ADM (blue) vs on-ADM (gray) in the placebo mITT set; per-visit sample sizes are shown below each panel. Forest plots of adjusted Δ values are shown in **Supplemental Figure 2**. Asterisks denote the PSW-adjusted LS mean difference (off-ADM minus on-ADM) at each visit: *P<0.05; **P<0.01; ***P<0.001; ****P<0.0001. *Note:* “Decline” is negative for cUHDRS, TFC, SWR, SDMT; positive for TMS (higher=worse).

**Supplemental Figure 2. PSW-adjusted forest plots of visit-wise differences (off-ADM vs on-ADM) across clinical outcomes (95% CI).** PSW estimates of LS mean change from baseline with 95% CI at Weeks 26, 39, 52, 65, and 78 for cUHDRS, TFC, SWR, SDMT, and TMS in the placebo mITT set; per-row n values are shown. Positive values indicate less worsening off-ADM for cUHDRS, TFC, SWR, and SDMT; for TMS (higher=worse), negative LS mean difference values favor off-ADM. P-values are listed at right.

**Supplemental Figure 3. TMS subdomains for VMAT2 inhibitor–only vs off-ADM (95% CI) forest plots.** LS mean difference (Δ) in change from baseline with 95% CI at Weeks 26, 39, 52, 65, and 78 for TMS subdomains: chorea, gait & balance, hand movements, bradykinesia, rigidity, eye movements, dystonia, and tongue protrusion. Per-row n and p-values are shown. For TMS negative LS mean differences favor off-ADM.

**Supplemental Figure 4. TMS subdomains with antipsychotic-only vs off-ADM (95% CI) forest plots.** LS mean difference (Δ) in change from baseline with 95% CI at Weeks 26, 39, 52, 65, and 78 for TMS subdomains (chorea, gait & balance, hand movements, bradykinesia, rigidity, eye movements, dystonia, tongue protrusion); per-row n and p-values shown. For TMS negative LS mean differences favor off-ADM.

**Supplemental Figure 5. VMAT2 inhibitor–only vs off-ADM across outcomes (95% CI) forest plots.** LS mean difference (Δ) in change from baseline with 95% CI for cUHDRS, TFC, SWR, SDMT, and TMS at Weeks 26, 39, 52, 65, and 78; per-row n values and p-values shown. Positive values favor off-ADM for cUHDRS, TFC, SWR, and SDMT; for TMS (higher=worse), negative LS mean difference values favor off-ADM.

**Supplemental Figure 6. Antipsychotic-only vs off-ADM across outcomes (95% CI) forest plots.** LS mean difference (Δ) in change from baseline with 95% CI at Weeks 26, 39, 52, 65, and 78 for cUHDRS, TFC, SWR, SDMT, and TMS in the placebo mITT set; per-row n values and p-values are shown. Positive differences favor off-ADM for cUHDRS, TFC, SWR, and SDMT; for TMS (higher=worse), LS mean negative differences favor off-ADM.

## Acknowledgements

The study was sponsored and funded by Prilenia Therapeutics, which supported the study design, trial oversight and conduct, site operations, data collection, statistical analysis, and manuscript development. The sponsor had final responsibility for verifying the accuracy of the data. We thank the participants in PROOF-HD and their families and caregivers. We acknowledge the contributions of site staff, sponsor teams, and the Huntington Study Group (HSG®) for coordination and support. We especially recognize the site principal investigators and study coordinators whose efforts were essential to parent trial execution. Programming support was provided by Tigermed India Clinical Research Pvt. Ltd. All authors contributed to drafting and revising this manuscript and approved the final version.

## Author Contributions and Financial Disclosures

**Karen Elta Anderson**

Contributed clinical interpretation, insight, and editorial support.

*Financial Disclosures:* Scientific Advisor to Prilenia, Site Investigator for PROOF-HD study, Site Investigator for PRIDE-HD.

**Andrew M. Tan**

Contributed to clinical data analysis and interpretation; drafted the manuscript and supervised its final production and submission with all authors.

*Financial Disclosures:* Employee of Prilenia Therapeutics B.V. with stock options.

**Andrew Feigin**

Served as the North American Principal Investigator; led clinical oversight and coordination in PROOF-HD (North America).

*Financial Disclosures:* Received grant support from Prilenia through NYU for his role as principal investigator for North American study sites.

**Ralf Reilmann**

Conceived and designed the study; served as European Principal Investigator; led clinical oversight and coordination in PROOF-HD (Europe).

*Financial Disclosures:* Founding director/owner of the George-Huntington-Institute (GHI) and QuantiMedis; has provided consulting, advisory, clinical trial operations, and Q-Motor analyses for Prilenia; principal investigator for European PROOF-HD sites; European coordinating investigator for PRIDE-HD.

**Anne E. Rosser**

Conceived and designed the study; served as European co-lead; led clinical oversight and coordination in PROOF-HD (Europe).

*Financial Disclosures:* Served as European co-lead for the PROOF-HD study.

**Lynn A. Raymond**

Contributed clinical interpretation, insight, and editorial support.

*Financial Disclosures:* None declared.

**Sandra K. Kostyk**

Served as North American co-lead; led clinical oversight and coordination in PROOF-HD (North America). Contributed clinical interpretation, insight, and editorial support.

*Financial Disclosures:* Medical Director of the HDSA Center of Excellence at The Ohio State University; Co-Chair, Huntington Study Group Executive Membership Committee; North American Co-PI of PROOF-HD.

**Carsten Saft**

Contributed clinical interpretation, insight, and editorial support.

*Financial Disclosures:* Declares no conflicts of interest related to this study.

**Kelly Chen**

Contributed statistical analysis and methodology, clinical data analysis and interpretation.

*Financial Disclosures:* Employee of Prilenia Therapeutics B.V. with stock options.

**Randal Hand**

Contributed to data interpretation; data analysis, and visual presentations in the manuscript.

*Financial Disclosures:* Employee of Prilenia Therapeutics B.V. with stock options.

**Michal Geva**

Conceived and designed the study; contributed to clinical data analysis and interpretation; editorial support in manuscript development; involved in funding acquisition and study sponsorship.

*Financial Disclosures:* Employee of Prilenia Therapeutics B.V. with stock options.

**Michael R. Hayden**

Conceived and designed the study; contributed to clinical data analysis and interpretation; involved in funding acquisition and study sponsorship. Supervised study and manuscript development.

*Financial Disclosures:* CEO and scientific co-founder of Prilenia Neurotherapeutics B.V.; Physician Scientist and University Killam Professor at the University of British Columbia; serves on the Boards of Ionis Pharmaceuticals, 89Bio, and AbCellera.

**This research was fully supported by Prilenia Therapeutics B.V. and/or its subsidiaries, which provided the necessary resources and funding.**

## Data Availability

De-identified participant data (IDP) with data dictionaries, along with key documents—the protocol, SAP, and informed-consent template—will be available to qualified researchers starting six months after publication and for five years thereafter.

Access is limited to investigators at academic or non-profit institutions pursuing scientifically sound, ethics-approved analyses that align with the study aims or address closely related questions. Because of ethical and legal constraints on participant privacy (e.g., GDPR), data are not placed in a public repository. Redacted versions of the PROOF-HD protocol and SAP can be accessed here: https://ghi-muenster.de/protocols/proof-hd and https://ghi-muenster.de/protocols/proof-hd-sap.

To request access, contact the Sponsor at. Requests are reviewed by the Sponsor or its data access committee, with a decision provided within 90 days. Approved users enter a Data Use Agreement that prohibits re-identification, disallows unauthorized data sharing, and limits use to the agreed purposes. Authorship or acknowledgment follows the International Committee of Medical Journal Editors (ICMJE) Recommendations (2024). A DUA template is available on request. No third-party proprietary datasets were used; all data were collected and analyzed by the investigators and the Sponsor as described in Methods.

## References

[1] Macdonald M. A novel gene containing a trinucleotide repeat that is expanded and unstable on Huntington’s disease chromosomes. Cell 1993; 72: 971–983.

[2] Ross CA, Aylward EH, Wild EJ, et al. Huntington disease: natural history, biomarkers and prospects for therapeutics. Nat Rev Neurol 2014; 10: 204–216.

[3] Marder K, Zhao H, Myers RH, et al. Rate of functional decline in Huntington’s disease. Huntington Study Group. Neurology 2000; 54: 452–458.

[4] Meltzer HY, McGurk SR. The Effects of Clozapine, Risperidone, and Olanzapine on Cognitive Function in Schizophrenia. Schizophr Bull 1999; 25: 233–256.

[5] Frank S. Tetrabenazine as anti-chorea therapy in Huntington disease: an open-label continuation study. Huntington Study Group/TETRA-HD Investigators. BMC Neurol 2009; 9: 62.

[6] Mestre T, Ferreira J, Coelho MM, et al. Therapeutic interventions for symptomatic treatment in Huntington’s disease. Cochrane Database Syst Rev 2009; CD006456.

[7] Armstrong MJ, Miyasaki JM, American Academy of Neurology. Evidence-based guideline: pharmacologic treatment of chorea in Huntington disease: report of the guideline development subcommittee of the American Academy of Neurology. Neurology 2012; 79: 597–603.

[8] Bashir H, Jankovic J. Treatment options for chorea. Expert Rev Neurother 2018; 18: 51–63.

[9] Siafis S, Wu H, Wang D, et al. Antipsychotic dose, dopamine D2 receptor occupancy and extrapyramidal side-effects: a systematic review and dose-response meta-analysis. Mol Psychiatry 2023; 28: 3267–3277.

[10] Zeng J, Raballo A, Ye J, et al. The impact of aripiprazole on neurocognitive function in individuals at clinical high risk for psychosis: A comparison with olanzapine and non-antipsychotic treatment. Eur Psychiatry J Assoc Eur Psychiatr 2025; 68: e69.

[11] Dean M, Sung V. Review of deutetrabenazine: a novel treatment for chorea associated with Huntington’s disease. Drug Des Devel Ther 2018; Volume 12: 313–319.

[12] Anderson K, Craufurd D, Edmondson MC, et al. An International Survey-based Algorithm for the Pharmacologic Treatment of Obsessive-Compulsive Behaviors in Huntington’s Disease. PLoS Curr 2011; 3: RRN1261.

[13] Anderson KE, Van Duijn E, Craufurd D, et al. Clinical Management of Neuropsychiatric Symptoms of Huntington Disease: Expert-Based Consensus Guidelines on Agitation, Anxiety, Apathy, Psychosis and Sleep Disorders. J Huntingt Dis 2018; 7: 355–366.

[14] Groves M, Van Duijn E, Anderson K, et al. An International Survey-based Algorithm for the Pharmacologic Treatment of Irritability in Huntington’s Disease. PLoS Curr 2011; 3: RRN1259.

[15] Achenbach J, Stodt B, Saft C. Factors Influencing the Total Functional Capacity Score as a Critical Endpoint in Huntington’s Disease Research. Biomedicines 2023; 11: 3336.

[16] Geva M, Goldberg YP, Schuring H, et al. Antidopaminergic Medications and Clinical Changes in Measures of Huntington’s Disease: A Causal Analysis. Mov Disord 2025; 40: 928–937.

[17] Trundell D, Palermo G, Schobel S, et al. Defining Clinically Meaningful Change on the Composite Unified Huntington’s Disease Rating Scale (cUHDRS) (P1.8-043). Neurology 2019; 92: P1.8–043.

[18] Geva M, Goldberg YP, Schuring H, et al. Antidopaminergic Medications and Clinical Changes in Measures of Huntington’s Disease: A Causal Analysis. Mov Disord.

[19] Boareto M, Yamamoto Y, Long JD, et al. Modeling Disease Progression and Placebo Response in Huntington Disease: Insights From Enroll-HD and GENERATION HD1 Cohorts. Neurology 2025; 104: e213646.

[20] Tedroff J, Waters S, Barker RA, et al. Antidopaminergic Medication is Associated with More Rapidly Progressive Huntington’s Disease. J Huntingt Dis 2015; 4: 131–140.

[21] Harris KL, Kuan W-L, Mason SL, et al. Antidopaminergic treatment is associated with reduced chorea and irritability but impaired cognition in Huntington’s disease (Enroll-HD). J Neurol Neurosurg Psychiatry 2020; 91: 622–630.

[22] Just KS, Dormann H, Freitag M, et al. CYP2D6 in the Brain: Potential Impact on Adverse Drug Reactions in the Central Nervous System—Results From the ADRED Study. Front Pharmacol; 12. Epub ahead of print 7 May 2021. DOI: 10.3389/fphar.2021.624104.

[23] Wang X, Huang J, Lu J, et al. Risperidone plasma level, and its correlation with CYP2D6 gene polymorphism, clinical response and side effects in chronic schizophrenia patients. BMC Psychiatry 2024; 24: 41.

[24] Tan AM, Geva M, Goldberg YP, et al. Antidopaminergic medications in Huntington’s disease. J Huntingt Dis 2025; 18796397241304312.

[25] Ghazaleh N, Houghton R, Palermo G, et al. Ranking the Predictive Power of Clinical and Biological Features Associated With Disease Progression in Huntington’s Disease. Front Neurol 2021; 12: 678484.

[26] Huntington Study Group. Tetrabenazine as antichorea therapy in Huntington disease: A randomized controlled trial. Neurology 2006; 66: 366–372.

[27] Reilmann R, Feigin A, Rosser AE, et al. Pridopidine in early-stage manifest Huntington’s disease: a phase 3 trial. Nat Med. Epub ahead of print 5 September 2025. DOI: 10.1038/s41591-025-03920-3.

[28] Kapur S, Remington G, Zipursky RB, et al. The D2 dopamine receptor occupancy of risperidone and its relationship to extrapyramidal symptoms: A pet study. Life Sci 1995; 57: PL103–PL107.

[29] Van Der Zwaan KF, Feleus S, Dekkers OM, et al. Total functioning capacity scale in Huntington’s disease: natural course over time. J Neurol 2025; 272: 140.

[30] Schobel SA, Palermo G, Auinger P, et al. Motor, cognitive, and functional declines contribute to a single progressive factor in early HD. Neurology 2017; 89: 2495–2502.

[31] Tiapride SmPC. Tiapride SUMMARY OF PRODUCT CHARACTERISTICS.

[32] package insert quetiapine. Seroquel (quetiapine) [package insert]. Astrazeneca.pdf.

[33] package insert tetrabenazine. Xenazine (tetrabenazine) [package insert]. Lundbeck.

[34] package insert deutetrabenazine. Austedo (deutetrabenazine) [package insert]. Teva Pharmaceuticals USA, Inc., https://www.accessdata.fda.gov/drugsatfda_docs/label/2017/208082s000lbl.pdf (2017).

[35] package insert. Aripiprazole (Abilify Asimtufii) [package insert]. Otsuka America Pharmaceutical, Inc..pdf.

[36] package insert risperidone. Risperdal (risperidone) [package insert]. Janssen Pharmaceuticals, Inc.

[37] Torrisi SA, Laudani S, Contarini G, et al. Dopamine, Cognitive Impairments and Second-Generation Antipsychotics: From Mechanistic Advances to More Personalized Treatments. Pharmaceuticals 2020; 13: 365.

[38] Bonelli RM, Niederwieser G, Tribl GG, et al. High-dose olanzapine in Huntington’s disease: Int Clin Psychopharmacol 2002; 17: 91.

[39] Ferreira JJ, Rodrigues FB, Duarte GS, et al. An Evidence Based Review on Treatments for Huntington’s Disease. Mov Disord 2022; 37: 25–35.

[40] Kapur S, Zipursky RB, Remington G, et al. 5-HT2 and D2 Receptor Occupancy of Olanzapine in Schizophrenia: A PET Investigation. Am J Psychiatry 1998; 155: 921–928.

[41] Ballard C, Howard R. Neuroleptic drugs in dementia: benefits and harm. Nat Rev Neurosci 2006; 7: 492–500.

[42] Ishihara L, Oliveri D, Wild EJ. Neuropsychiatric comorbidities in Huntington’s and Parkinson’s Disease: A United States claims database analysis. Ann Clin Transl Neurol 2021; 8: 126–137.

[43] Andriessen RL, Oosterloo M, Hollands A, et al. Psychotropic medication use in Huntington’s disease: A retrospective cohort study. Parkinsonism Relat Disord 2022; 105: 69–74.

[44] Orth M, Bronzova J, Tritsch C, et al. Comparison of Huntington’s Disease in Europe and North America. Mov Disord Clin Pract 2016; 4: 358–367.

[45] Lock SK, Kappel DB, Owen MJ, et al. Antipsychotic and pharmacogenomic effects on cross-sectional symptom severity and cognitive ability in schizophrenia. eBioMedicine 2025; 116: 105745.

[46] Dickerson F, Origoni A, Katsafanas E, et al. The Association Between Psychotropic Medications and Cognitive Functioning in a Real-World Cohort of 869 Individuals with Schizophrenia. Schizophr Bull. Epub ahead of print 8 May 2025. DOI: 10.1093/schbul/sbaf047.

