## Supplementary material for "Antidopaminergic Medications Are Associated with Faster Decline in Measures of Clinical Outcome in HD: Insights from PROOF-HD": Dose thresholds for VMAT2 inhibitors and antipsychotics used for dose response subgrouping

|  | **Lower Dose Cutoff (mg/day)** | **Higher Dose Cutoff (mg/day)** |
| --- | --- | --- |
| **VMAT2 inhibitors (not on antipsychotics)** | | |
| **Deutetrabenazine (dTBZ)** | < 30, n=6 | ≥ 30, n=11 |
| **Tetrabenazine (TBZ)** | ≤ 50, n=8 | > 50, n=4 |
| **VMAT2 inhibitors (TBZ, dTBZ)** | N=14 | N=15 |
| **Antipsychotics (not on VMAT2i)** | | |
| **Aripiprazole** | ≤ 5, n=3 | > 5, n=10 |
| **Quetiapine** | ≤ 50, n=2 | > 50, n=10 |
| **Olanzapine** | ≤ 5, n=9 | > 5, n=18 |
| **Risperidone** | ≤ 0.5, n=5 | > 0.5, n=21 |

### Supplemental Table 1
