## Supplementary material for "Antidopaminergic Medications Are Associated with Faster Decline in Measures of Clinical Outcome in HD: Insights from PROOF-HD": ADM exposure frequencies in the placebo arm (mITT).

|  | PROOF-HD  Placebo arm, mITT  Total (on and off ADMs) N=247 |
| --- | --- |
| Off ADMs all the time during the study | 112 (45.3%) |
| On ADMs anytime during the study | 135 (54.7%) |
| On at least 1 antipsychotic anytime during the study (not on VMAT2i) | 87 (35.2%) |
| On VMAT2i anytime during the study (not on antipsychotics) | 29 (11.7%) |
| *On >1 ADM anytime during the study | 19 (7.7%) |

### Supplemental Table 2
