## Supplementary material for "Antidopaminergic Medications Are Associated with Faster Decline in Measures of Clinical Outcome in HD: Insights from PROOF-HD": Antipsychotic and VMAT2i use by agent (non mutually exclusive).

| **Drug** | **Total population, mITT**  **(On-ADM and Off-ADM, n=247)** |
| --- | --- |
| **Antipsychotics**  *Note: Patients receiving only antipsychotic medications may be prescribed multiple antipsychotics and therefore may be counted in more than one row* | |
| Olanzapine | 27 (10.9%) |
| Risperidone | 26 (10.5%) |
| Tiapride | 18 (7.3%) |
| Aripiprazole | 13 (5.3%) |
| Quetiapine | 12 (4.9%) |
| Other (e.g., fluphenazine, haloperidol) | 13 (5.3%) |
| **VMAT2 Inhibitors** | |
| Deutetrabenazine | 17 (6.9%) |
| Tetrabenazine | 12 (4.9%) |

### Supplemental Table 3
