## Supplementary material for "Antidopaminergic Medications Are Associated with Faster Decline in Measures of Clinical Outcome in HD: Insights from PROOF-HD": Percent (%) decline off vs on-ADMs (unadjusted and PSW).

### Supplemental Table 4

|  | **cUHDRS** | | **TFC** | | **SWR** | | **SDMT** | | **TMS** | |
| --- | --- | --- | --- | --- | --- | --- | --- | --- | --- | --- |
| Timepoint | **(Unadj)** | **(PSW)** | **(Unadj)** | **(PSW)** | **(Unadj)** | **(PSW)** | **(Unadj)** | **(PSW)** | **(Unadj)** | **(PSW)** |
| **Week 26** | 46.1 | 42.0 | 66.0 | 44.9 | 83.6 | 80.1 | 81.2 | 18.1 | 136.5 | 125.0 |
| **Week 39** | 62.4 | 53.2 | 66.6 | 49.0 | 95.8 | 87.4 | 98.3 | 14.8 | 77.4 | 43.8 |
| **Week 52** | 70.5 | 64.8 | 77.6 | 69.7 | 99.3 | 69.0 | 95.7 | 88.0 | 75.0 | 75.6 |
| **Week 65** | 62.6 | 57.8 | 59.4 | 58.6 | 91.8 | 90.0 | 117.4 | 120.3 | 63.8 | 25.7 |
| **Week 78** | 69.4 | 52.2 | 63.6 | 54.4 | 91.6 | 39.2 | 117.4 | 92.6 | 64.9 | 39.0 |
