## Supplementary figures and images for "Antidopaminergic Medications Are Associated with Faster Decline in Measures of Clinical Outcome in HD: Insights from PROOF-HD"

### Antipsychotic only vs off ADM across outcomes (95% CI) forest plots.

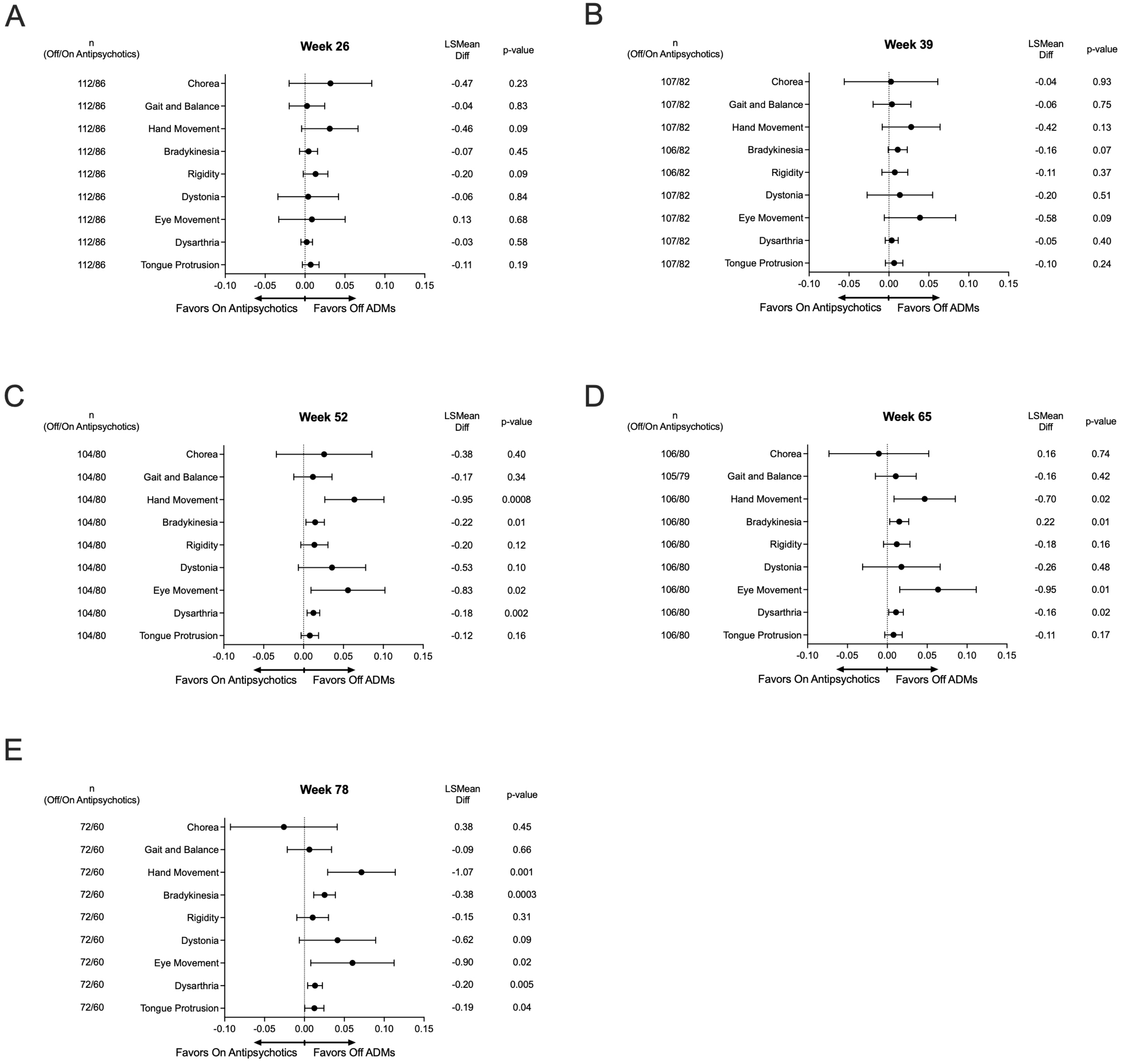

### Propensity score weighted clinical outcomes by ADM exposure.

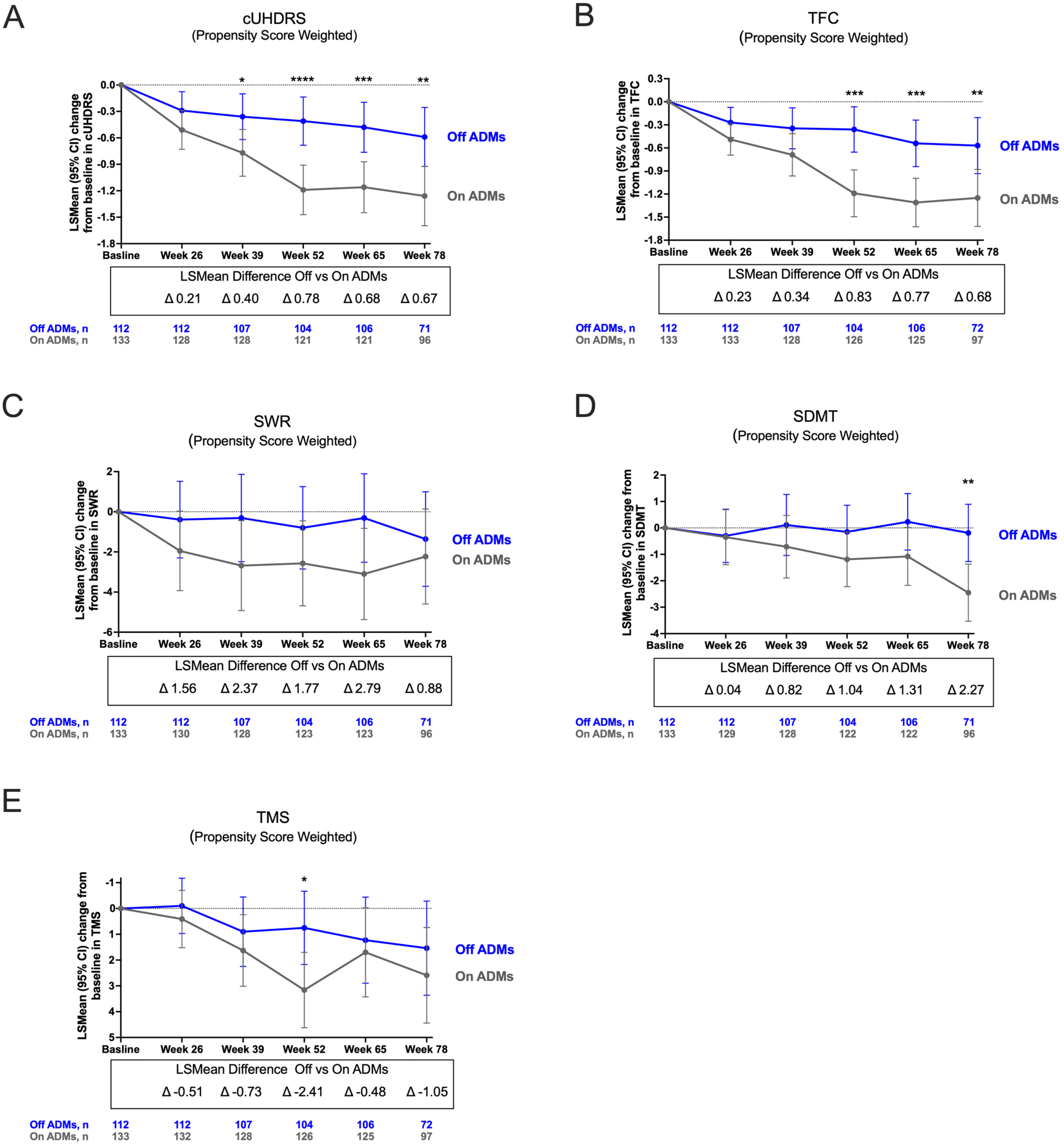

### PSW adjusted forest plots of visit-wise differences (off-ADM vs on-ADM) across clinical outcomes (95% CI).

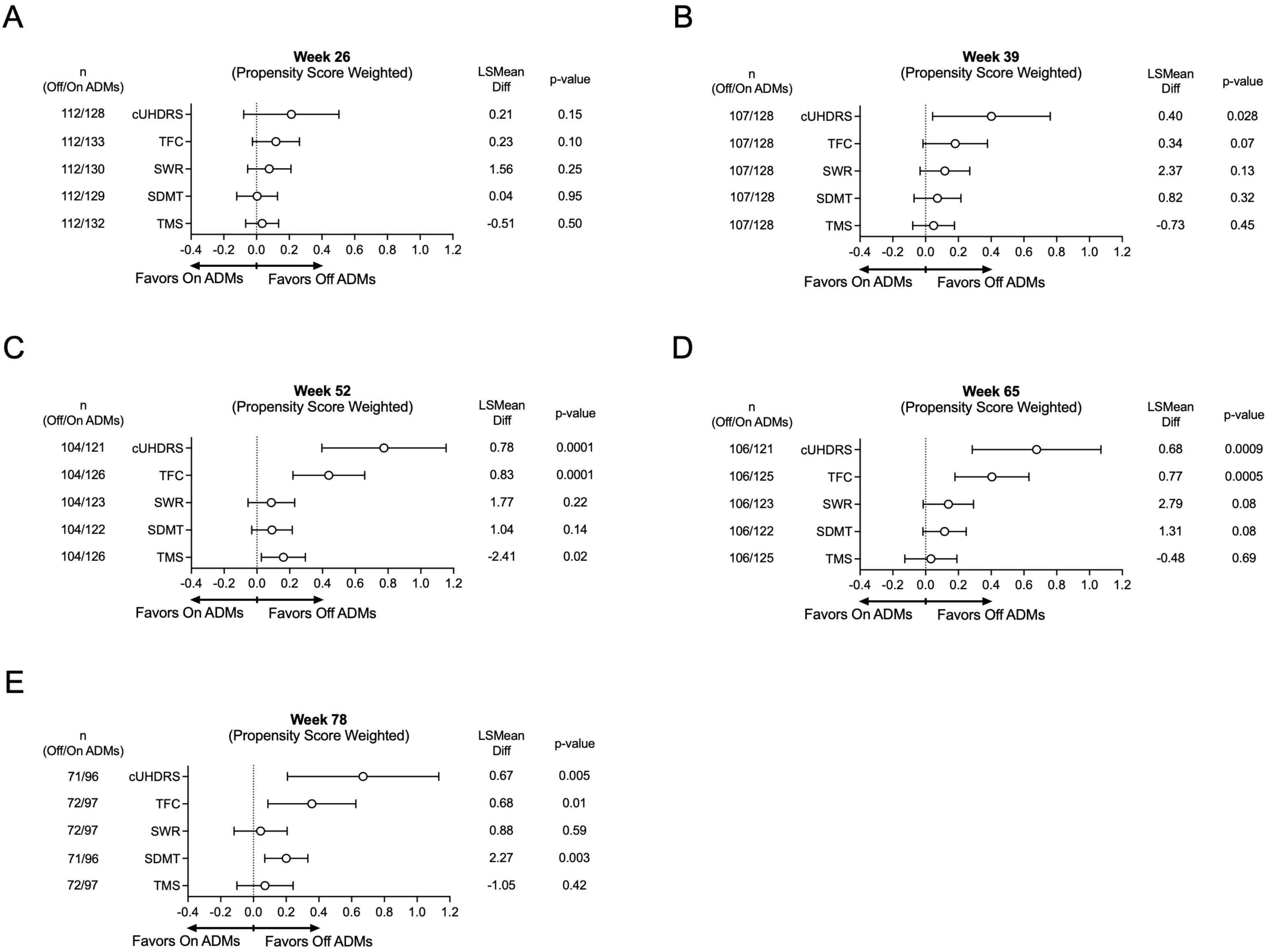

### TMS subdomains for VMAT2 inhibitor only vs off-ADM (95% CI) forest

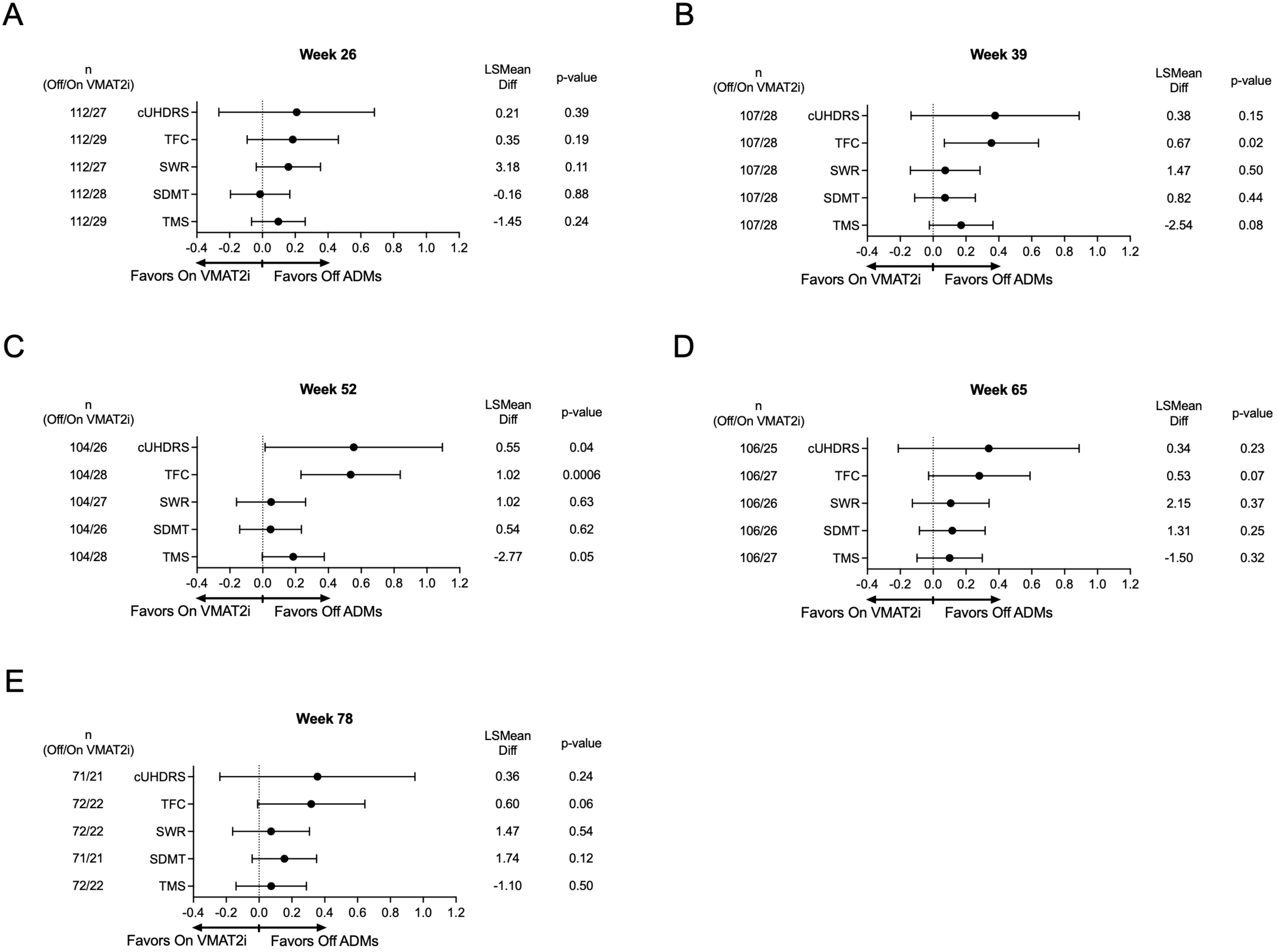

### TMS subdomains with antipsychotic only vs off-ADM (95% CI) forest plots.

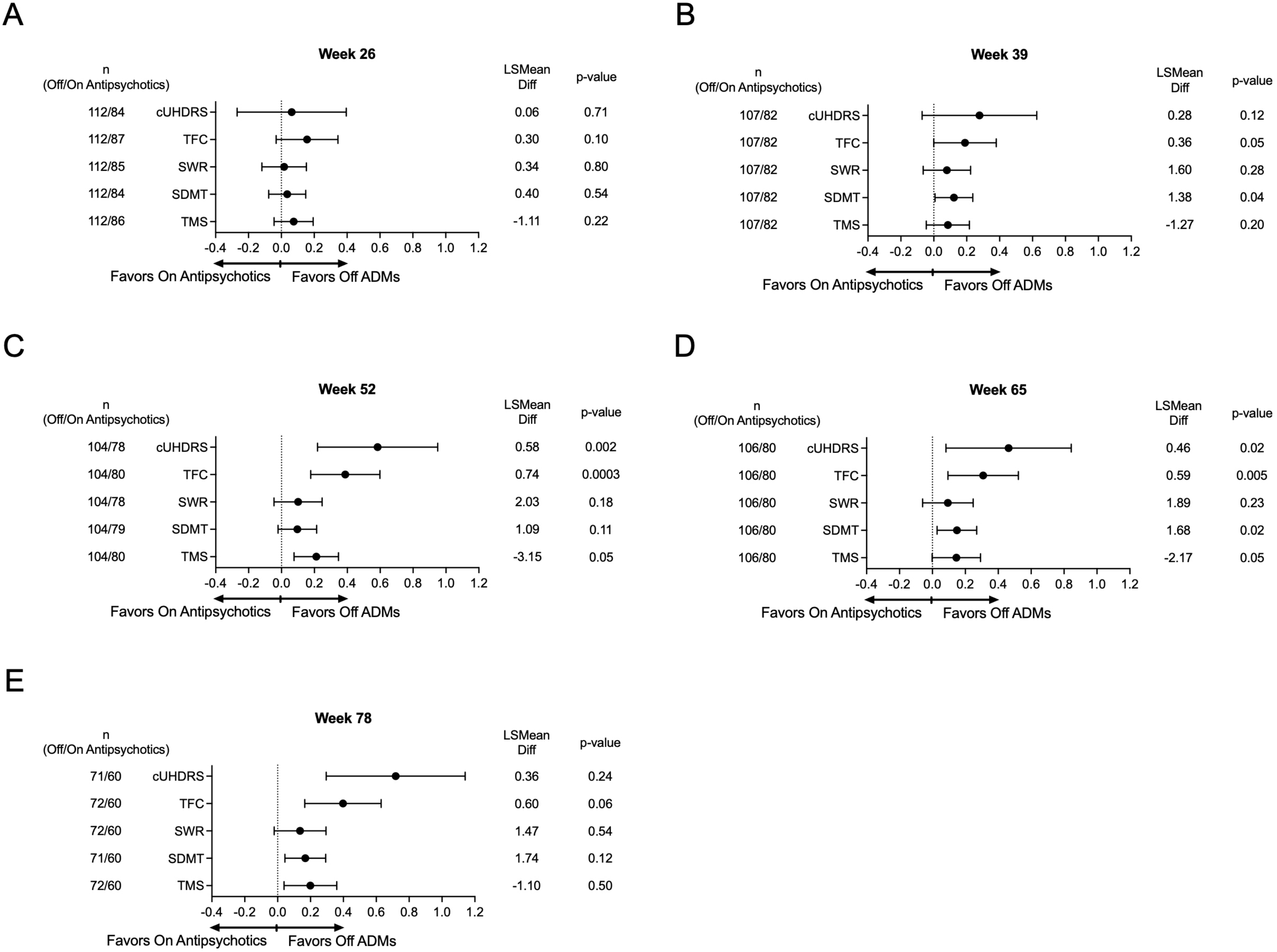

### VMAT2 inhibitor only vs off-ADM across outcomes (95% CI) forest plots.

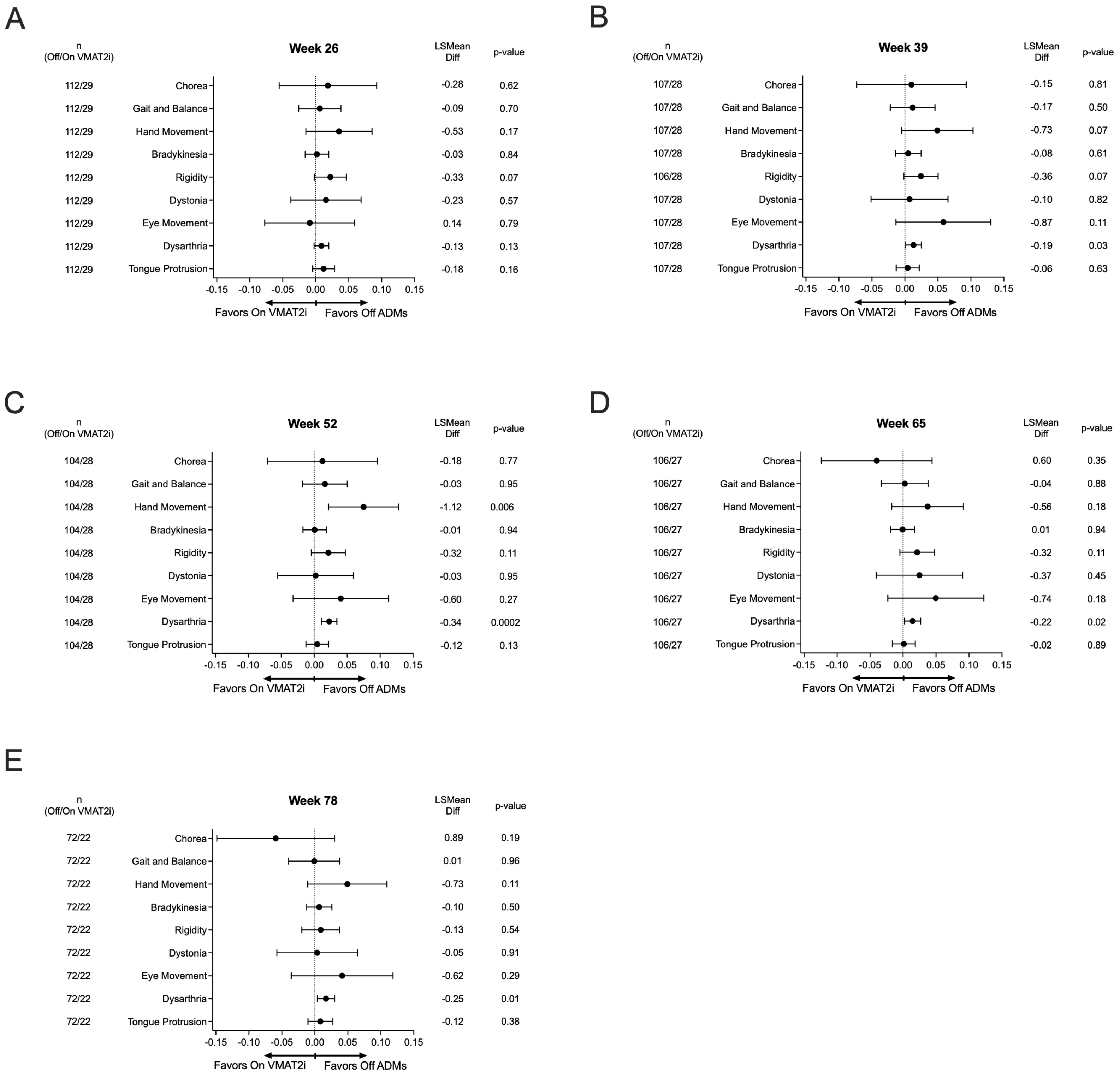
