## Supplemental Material (results extended text) for "Antidopaminergic Medications Are Associated with Faster Decline in Measures of Clinical Outcome in HD: Insights from PROOF-HD"

### Impact of ADM Use on Measures of Clinical Outcome

To evaluate whether ADM exposure influenced the longitudinal trajectory of clinical decline, we assessed change from baseline across clinical outcome measures in the placebo arm of the PROOF-HD trial (on-ADM vs off-ADM). Outcomes included cUHDRS, TFC, SDMT, SWR, and TMS (**Figure 1**).

In cUHDRS (**Figure 1A**), the on-ADM group demonstrated a faster decline as compared with the off-ADM group, with between-group differences emerging by Week 39 and persisting through study end. At Week 26, the LS mean change from baseline was –0.45 in the on-ADM group versus –0.30 in the off-ADM group (Δ=0.15; p=0.34, 95% CI −0.16 to 0.46). Differences became statistically significant at Week 39 (Δ=0.38; p=0.03, 95% CI 0.04 to 0.71), and increased in magnitude through Week 52 (Δ=0.66; p=0.0002, 95% CI 0.31 to 1.01) and Week 65 (Δ=0.58; p=0.002, 95% CI 0.22 to 0.94). **At Week 65, the percent reduction in decline off-ADMs as compared with on-ADMs was 62.6%, and by Week 78, further reduced by 69.1%.** Specifically, by Week 78, the between-group difference peaked (Δ=0.78; p=0.0002, 95% CI 0.38 to 1.19), with mean cUHDRS decline of –1.28 in the on-ADM group versus –0.49 in the off-ADM group. These results show a consistent association between ADM exposure and greater outcome worsening as measured by cUHDRS.

Similarly, as shown in TFC scores (**Figure 1B**), functional decline was observed in both groups but was consistently worse among participants on-ADM. At Week 26, the LS mean change was –0.57 in the on-ADM group and –0.21 in the off-ADM group (Δ=0.36; p=0.04, 95% CI 0.01 to 0.70). The difference widened at Week 39 (Δ=0.46; p=0.01, 95% CI 0.11 to 0.80) and reached its greatest magnitude at Week 52 (Δ=0.85; p<0.0001, 95% CI 0.47 to 1.22), remaining significant through Week 65 (Δ=0.66; p=0.0008, 95% CI 0.28 to 1.05) and Week 78 (Δ=0.84; p=0.0001, 95% CI 0.41 to 1.26), where mean TFC decline was –1.28 versus –0.45. At Week 65, the percent reduction in decline off-ADMs as compared with on-ADMs was 59.4%, and at Week 78 it was 67.6%.

In cognitive measures (SWR and SDMT; **Figure 1C** and **Figure 1D**), on-ADM participants demonstrated faster trajectories. For SWR (**Figure 1C**), mean change from baseline was –1.89 (on-ADM) and –0.38 (off-ADM) at Week 26 (Δ=1.51; p=0.21, 95% CI −0.87 to 3.89). Group differences increased at Week 39 (Δ=2.03; p=0.12, 95% CI −0.53 to 4.59) and Week 52 (Δ=2.28; p=0.08, 95% CI −0.31 to 4.86), reaching statistical significance by Week 65 (Δ=2.87; p=0.04, 95% CI 0.13 to 5.62) and Week 78 (Δ=3.26; p=0.03, 95% CI 0.35 to 6.17), with mean declines of –3.96 and –0.70, respectively. **At** Week 65, the percent reduction in decline off-ADMs as compared with on-ADMs was 91.8%, and at Week 78 it was 85.6%.

For SDMT (**Figure 1D**), the between-group difference was not significant at Week 26 (Δ=0.51; p=0.39, 95% CI −0.65 to 1.68) but became significant at Week 39 (Δ=1.60; p=0.0098, 95% CI 0.39 to 2.81). At Week 52, the trend persisted (Δ=1.17; p=0.06, 95% CI −0.03 to 2.37), with stronger effects at Week 65 (Δ=1.94; p=0.0026, 95% CI 0.68 to 3.20) and Week 78 (Δ=2.36; p=0.0004, 95% CI 1.07 to 3.66), where mean scores changed by –2.31 (on-ADM) versus +0.06 (off-ADM). At Week 65, the percent reduction in decline off-ADMs as compared with on-ADMs was 117.4%, and at Week 78 it was 102.0%.

Motor progression, as measured by TMS (**Figure 1E**), also differed between groups, with participants on-ADM showing faster motor decline. Note that for TMS, lower [negative] change indicates improvement. At Week 26, mean change was +0.76 in the on-ADM group and –0.24 in the off-ADM group (Δ=–1.00; p=0.24, 95% CI −2.65 to 0.66). Differences grew at Week 39 (Δ=–1.76; p=0.06, 95% CI −3.60 to 0.09), reaching statistical significance at Week 52 (Δ=–3.18; p=0.0010, 95% CI −5.07 to −1.30), Week 65 (Δ=–2.39; p=0.02, 95% CI −4.41 to −0.36), and Week 78 (Δ=–2.81; p=0.01, 95% CI −4.98 to −0.64). At study end, mean TMS change was +3.94 in the on-ADM group versus +1.13 in the off-ADM group. **At Week 65, the percent reduction in decline off-ADMs as compared with on-ADMs was 65.8%, and at Week 78 it was 66.9%.**

### Impact of ADM Use on Measures of Clinical Outcome (PSW Analyses)

To address possible residual baseline imbalance, we conducted a PSW analysis (cohort baseline data shown in **Table 1**). **Briefly, to PS-weight adjust for baseline differences between the on- and off-ADM groups in key metrics related to HD, average treatment effects in the treated (ATT) weights were estimated using a logistic model, with ADM use (off vs on) as the dependent variable, and following baseline covariates: age, sex, region, CAG repeats, CAP100 (calculated as age x (CAG-30)/6.49), TFC, TMS, SWR, SDMT, and Q-Motor FT-IOI mean. To mitigate the influence of extreme values, patients with propensity scores >95th percentile or <5th percentile were excluded. Two patients on-ADMs were excluded from the PSW population using this criterion. The resulting weights were then applied in an MMRM analysis consistent with the primary model, but restricted to the placebo arm and excluding the** treatment-by-visit and treatment-by-ADM use interaction terms.

In cUHDRS, PSW-adjusted analyses confirmed the pattern shown for the primary, unadjusted analyses, with significant separation at Week 39 (Δ=0.402; p=0.0284, 95% CI 0.043 to 0.761), Week 52 (Δ=0.775; p=0.0000868, 95% CI 0.396 to 1.154), Week 65 (Δ=0.677; p=0.00086, 95% CI 0.284 to 1.070), and Week 78 (Δ=0.670; p=0.0048, 95% CI 0.207 to 1.132) (**Supplemental** **Figure 1A**). The PSW percent reduction in decline off-ADMs as compared with on-ADMs was 57.8% at Week 65 and 52.2% at Week 78.

For TFC, PSW-adjusted analyses also followed the same pattern as the unadjusted outcome, with separation emerging by Week 26 (Δ=0.23; p=0.1046, 95% CI −0.05 to 0.50) and Week 39 (Δ=0.34; p=0.0711, 95% CI −0.03 to 0.72), becoming statistically significant at Week 52 (Δ=0.83; p=0.0001, 95% CI 0.42 to 1.25), Week 65 (Δ=0.77; p=0.0005, 95% CI 0.34 to 1.20), and Week 78 (Δ=0.68; p=0.0095, 95% CI 0.17 to 1.19) (**Supplemental** **Figure 1B**). The PSW percent reduction in decline off-ADMs as compared with on-ADMs was 58.8% at Week 65 and 54.4% at Week 78.

For SWR, PSW-adjusted analyses showed the same late-visit separation, with significant differences at Week 65 (Δ=2.79; p=0.0761, 95% CI −0.3 to 5.88) and Week 78 (Δ=0.878; p=0.5944, 95% CI −2.38 to 4.13) (**Supplemental** **Figure 1C**). The PSW percent reduction in decline off-ADMs as compared with on-ADMs was 90.0% at Week 65 and 39.2% at Week 78.

In SDMT measures, PSW-adjusted results were similar, with significant differences at Week 39 (Δ=0.820; p=0.3166, 95% CI −0.78 to 2.42), Week 65 (Δ=1.309; p=0.0848, 95% CI −0.18 to 2.80), and Week 78 (Δ=2.2651; p=0.0031, 95% CI 0.76 to 3.77) (**Supplemental** **Figure 1D**). The PSW percent reduction in decline off-ADMs as compared with on-ADMs was 120.3% at Week 65 and 92.6% at Week 78.

For TMS, PSW-adjusted analyses (**Supplemental** **Figure 1E**) similarly showed greater motor worsening with ADM exposure, with significant difference at Week 52 (Δ=–2.405; p=0.0189, 95% CI −4.41 to −0.40). The difference at Week 65 (Δ=–0.4785; p=0.6907, 95% CI −2.85 to 1.89), and Week 78 (Δ=–1.0529; p=0.4180, 95% CI −3.60 to 1.50). The PSW percent reduction in decline off-ADMs as compared with on-ADMs was 25.7% at Week 65 and 39.0% at Week 78.

Overall, PSW confirmed the mid-to-late between-group separation seen in the unadjusted analyses (shown in **Figure 1**). Moreover, PSW results were directionally consistent with the unadjusted findings, with mid-to-late separation remaining significant for cUHDRS and TFC. Forest plots (**Supplemental Figure 2**) show the magnitude and precision of PSW-adjusted between-group differences across visits, with a consistent pattern favoring off-ADM over on-ADM for cUHDRS, TFC, SWR, SDMT, and TMS. Taken together, findings demonstrate that participants in off-ADMs experienced less decline throughout the double-blind trial period, supporting a consistent association between ADM exposure and faster disease progression.

#### Effect of ADM Exposure on TMS Subdomains

As VMAT2i and many antipsychotics are prescribed for symptom management of motor disturbances, particularly chorea, we evaluated the pattern of treatment effect of ADM exposure across individual TMS subdomains (**Figure 2**). Note that positive change values in motor outcome measures indicate worse outcomes. Although participants on-ADMs showed favorable TMS-chorea scores at later timepoints (Week 65: Δ=0.14, p=0.74, 95% CI −0.71 to 0.99; Week 78: Δ=0.42, p=0.37, 95% CI −0.49 to 1.33), these differences were not statistically significant. However, in other TMS subdomain measures, participants off-ADMs had consistently improved scores beginning by Week 52; especially with the largest, significant differences observed in Hand movements (Week 52: Δ=−1.13, p<0.0001, 95% CI −1.64 to −0.62; Week 65: Δ=−0.80, p=0.003, 95% CI −1.34 to −0.27; Week 78: Δ=−1.07, p=0.0004, 95% CI −1.66 to −0.48) and Eye movements (Week 65: Δ=−1.03, p=0.003, 95% CI −1.72 to −0.35; Week 78: Δ=−0.88, p=0.02, 95% CI −1.60 to −0.17). Dysarthria (Week 52: Δ=−0.23, p<0.0001; Week 65: Δ=−0.18, p=0.003; Week 78: Δ=−0.22, p=0.0008), Tongue protrusion reached significance at Week 78 (Δ=−0.18, p=0.042, 95% CI −0.361 to −0.0066), and Bradykinesia favored off-ADMs at Weeks 65 (Δ=−0.20, p=0.02, 95% CI −0.37 to −0.03) and Week 78 (Δ=−0.33, p=0.0006, 95% CI −0.52 to −0.14). Rigidity trended toward benefit without significance at Week 65 (Δ=−0.21, p=0.08, 95% CI −0.44 to 0.03) and Week 78 (Δ=−0.16, p=0.28, 95% CI −0.44 to 0.13).

**As shown in ADM** class-specific **forest plots,** VMAT2i-only participants (**Supplemental Figure 3**) show a small, nonsignificant tendency toward less chorea worsening, **with broader non-chorea declines** as compared with **off-ADM**. Antipsychotic-only comparisons (**Supplemental Figure 4**) show directionally similar **or more pronounced non-chorea** differences **favoring off-ADM, particularly** for bradykinesia and dysarthria. Overall, in a placebo-arm analysis of PROOF-HD, ADM exposure was associated with greater decline across multiple non-chorea TMS subdomains as compared with off-ADMs.

#### Impact of ADM Class on Clinical Progression: VMAT2 inhibitors and Antipsychotics

To examine potential class-specific associations between ADM exposure and clinical progression, we conducted exploratory analyses within the placebo arm. As shown in **Figure 3,** participants on VMAT2i-only or antipsychotic-only were compared with off-ADM participants (i.e., not exposed to any of these drugs).

#### Effect of VMAT2 Inhibitors on Clinical Outcomes

In cUHDRS (**Figure 3A**), both groups declined over time. Separation favoring off-ADMs was apparent and reached nominal significance at Week 52 (Δ=0.554; 95% CI −1.094 to −0.014; p=0.044). Earlier visits showed a similar, non-significant trend (Week 26: Δ=0.209; 95% CI −0.683 to 0.266; p=0.387; Week 39: Δ=0.378; 95% CI −0.889 to 0.134; p=0.148). Later visits were likewise not significant (Week 65: Δ=0.338; 95% CI −0.888 to 0.212; p=0.228; Week 78: Δ=0.356; 95% CI −0.950 to 0.239; p=0.240).

TFC showed a similar pattern (**Figure 3B**), with significant differences favoring off-ADMs at Week 39 (Δ=0.675; 95% CI −1.219 to −0.130; p=0.015) and Week 52 (Δ=1.017; 95% CI −1.592 to −0.442; p=0.00057). Earlier and later visits were directionally consistent without significance (Week 26: Δ=0.351; 95% CI −0.879 to 0.177; p=0.191; Week 65: Δ=0.534; 95% CI −1.120 to 0.052; p=0.074; Week 78: Δ=0.604; 95% CI −1.225 to 0.017; p=0.057).

Cognitive outcomes were directionally aligned with less decline off-ADM but did not reach significance (**Figure 3C–D**). For SWR, differences favored off-ADM at all visits without achieving significance: Week 26 Δ=3.18 (95% CI −7.12 to 0.75; p=0.112), Week 39 Δ=1.47 (−5.74 to 2.79; p=0.497), Week 52 Δ=1.02 (−5.25 to 3.20; p=0.634), Week 65 Δ=2.15 (−6.84 to 2.54; p=0.367), and Week 78 Δ=1.47 (−6.17 to 3.24; p=0.540). SDMT showed a similar overall pattern, with a small, non-significant difference at Week 26 Δ=0.16 (−1.88 to 2.20; p=0.878) that shifted in the off-ADM direction at later visits: Week 39 Δ=0.80 (−2.90 to 1.27; p=0.441), Week 52 Δ=0.54 (−2.66 to 1.59; p=0.621), Week 65 Δ=1.32 (−3.58 to 0.95; p=0.253), and Week 78 Δ=1.74 (−3.96 to 0.48; p=0.123).

Motor function as measured by TMS (**Figure 3E**) also favored the off-ADM group across visits but was not significant (Week 26: Δ=1.45, 95% CI −0.99 to 3.89; p=0.244; Week 39: Δ=2.54, 95% CI −0.35 to 5.43; p=0.085; Week 52: Δ=2.77, 95% CI −0.05 to 5.58; p=0.054; Week 65: Δ=1.50, 95% CI −1.47 to 4.46; p=0.322; Week 78: Δ=1.10, 95% CI −2.09 to 4.30; p=0.498).

To illustrate the directional changes between cohorts exposed to VMAT2 inhibitors-only versus off-ADM participants (**Supplemental Figure 5**), forest plots show a consistent pattern favoring off-ADM after Week 26 across cUHDRS, TFC, SWR, SDMT, and TMS, with the largest separations between Weeks 39 and 52. Taken together, these exploratory findings indicate less decline off-ADM across the double-blind period, consistent with an association between VMAT2i exposure and faster progression.

#### Effect of Antipsychotics on Clinical Outcomes

In cUHDRS, scores revealed greater clinical decline among participants receiving antipsychotics **as compared with** those who remained off ADMs (**Figure 3A**). By Week 26, the LS mean difference (off vs on antipsychotics) was 0.06 (95% CI −0.40 to 0.27; p=0.709), indicating minimal separation. A similar trend is shown at Week 39 (Δ=0.28; 95% CI −0.63 to 0.07; p=0.119). The difference favoring off-ADM reached significance at Week 52 (Δ=0.58; 95% CI 0.22 to 0.95; p=0.0018) and remained significant through the end of the study: Week 65 (Δ=0.46; 95% CI 0.08 to 0.84; p=0.017) and Week 78 (Δ=0.72; 95% CI 0.30 to 1.14; p=0.00091).

TFC also declined more rapidly among **participants** on antipsychotics than those off ADMs (**Figure 3B**). By Week 26, the between-group difference in LS mean change from baseline (off vs on antipsychotics) was 0.30 (95% CI −0.06 to 0.65; p=0.103), and was numerically larger at Week 39 (Δ=0.36; 95% CI −0.00 to 0.72; p=0.051). At later visits, scores were significant: Week 52 (Δ=0.74; 95% CI 0.34 to 1.14; p=0.0003), Week 65 (Δ=0.59; 95% CI 0.18 to 0.99; p=0.0049), and Week 78 (Δ=0.76; 95% CI 0.31 to 1.20; p=0.0008).

Antipsychotic exposure was associated with greater cognitive decline on SDMT, with directionally similar but non-significant differences on SWR (**Figure 3C–D**). For SWR, between-group differences favored the off-ADM group at all visits but did not reach significance (Week 26: Δ Week =0.34; 95% CI −2.38 to 3.06; p=0.805; 39: Δ=1.60; 95% CI −1.30 to 4.50; p=0.279; Week 52: Δ=2.03; 95% CI −0.92 to 4.97; p=0.177; Week 65: Δ=1.89; 95% CI −1.20 to 4.98; p=0.230; Week 78: Δ=2.74; 95% CI −0.43 to 5.91; p=0.090). In contrast, SDMT showed significant separation at multiple timepoints; at Week 39 (Δ=1.38; 95% CI 0.09 to 2.68; p=0.037), Week 65 (Δ=1.68; 95% CI 0.32 to 3.04; p=0.015), and Week 78 (Δ=1.90; 95% CI 0.50 to 3.31; p=0.008), with non-significant differences at Week 26 (Δ=0.40; 95% CI −0.87 to 1.67; p=0.535) and Week 52 (Δ=1.09; 95% CI −0.24 to 2.42; p=0.107), indicating a consistent association between antipsychotic exposure and worse processing speed.

**Motor progression** was also faster in participants receiving antipsychotics, with significantly higher (worse) TMS scores at several timepoints **as compared with** those off ADMs (**Figure 3E**). The between-group difference reached statistical significance at Week 52 (Δ=−3.15; 95% CI −5.16 to −1.14; p=0.0022) and Week 78 (Δ=−2.97; 95% CI −5.36 to −0.58; p=0.015), with a trending effect at Week 65 (Δ=−2.17; 95% CI −4.36 to 0.02; p=0.052). For completeness, earlier visits were not significant: Week 26 (Δ=−1.11; 95% CI −2.89 to 0.66; p=0.218) and Week 39 (Δ=−1.27; 95% CI −3.22 to 0.68; p=0.200).

As shown in **Supplemental Figure 6**, antipsychotic-only participants showed greater worsening than off-ADM across cUHDRS, TFC, SDMT, and TMS, with the most pronounced separations later in the trial post-Week 26 which persisted to study end.

Taken together, in these comparisons both VMAT2 inhibitors- and antipsychotic-only exposure were associated with greater decline on in global and functional outcomes as compared with off-ADMs. **Statistically robust differences versus off-ADM were observed for antipsychotic-only across TFC, cUHDRS, SDMT, and TMS, while VMAT2i showed similar directional trends. Note that** direct class-to-class comparisons were not performed, due in part to smaller on-ADM cohorts which limited statistical power and thus also warrant cautious interpretation of these findings.

#### Dose-Dependent Effects of ADMs on Clinical Outcomes

**ADM effects are dose-dependent** [1–5]**.** As shown in **Figure 4**, participants on **higher-dose** or **lower-dose** ADMs were compared with off-ADM across cUHDRS, TFC, SWR, SDMT, and TMS (i.e, tetrabenazine, deutetrabenazine, quetiapine, aripiprazole, risperidone, olanzapine; dose thresholds shown in **Supplemental Table 1**).

In **cUHDRS**, higher-dose ADMs significantly separated by Week 39 and maintained worsened measures until the end of the study: Week 26 Δ=0.1565 (95% CI −0.2109 to 0.5238; p=0.4030), Week 39 Δ=0.3979 (95% CI 0.0091 to 0.7867; p=0.0449), Week 52 Δ=0.7740 (95% CI 0.3599 to 1.1880; p=0.0003), Week 65 Δ=0.7072 (95% CI 0.2814 to 1.1329; p=0.0012), Week 78 Δ=0.9961 (95% CI 0.5257 to 1.4666; p=<0.0001), each as compared with off-ADM (Figure 4A). In contrast, lower-dose ADMs remained similar to the off-ADM without detectable significance at any visit: Week 26 Δ=0.1704 (95% CI −0.2945 to 0.6352; p=0.4717), Week 39 Δ=0.1750 (95% CI −0.3154 to 0.6655; p=0.4835), Week 52 Δ=0.4056 (95% CI −0.1142 to 0.9255; p=0.1259), Week 65 Δ=0.2876 (95% CI −0.2466 to 0.8218; p=0.2908), Week 78 Δ=0.2040 (95% CI −0.3852 to 0.7931; p=0.4967).

TFC measures followed a similar pattern with higher-dose ADMs showing differences that were significant as compared with off-ADM that appeared early in the study and maintained at every visit: Week 26 Δ=0.6176 (95% CI 0.2194 to 1.0159; p=0.0024), Week 39 Δ=0.7028 (95% CI 0.3002 to 1.1054; p=0.0007), Week 52 Δ=1.2048 (95% CI 0.7735 to 1.6361; p=<0.0001), Week 65 Δ=1.0612 (95% CI 0.6176 to 1.5048; p=<0.0001), Week 78 Δ=1.2790 (95% CI 0.7919 to 1.7661; p=<0.0001) (**Figure 4B**). With lower-dose ADMs, effect sizes remained relatively small and non-significant similar across all visits as compared with off ADMs: Week 26 Δ=0.1610 (95% CI −0.3528 to 0.6748; p=0.5384), Week 39 Δ=0.1271 (95% CI −0.3914 to 0.6455; p=0.6303), Week 52 Δ=0.3812 (95% CI −0.1709 to 0.9333; p=0.1756), Week 65 Δ=0.1148 (95% CI −0.4522 to 0.6818; p=0.6910), Week 78 Δ=0.0785 (95% CI −0.5419 to 0.6989; p=0.8037).

Both cognitive measures, SWR and SDMT, reached significant differences only later in the trial with higher dose as compared with the off-ADM cohort. For SWR, higher-dose ADMs showed a worsening trend as compared with the off-ADM group from early visits (Week 26 Δ=0.8722, 95% CI −1.9305 to 3.6750; p=0.5412; Week 39 Δ=1.5108, 95% CI −1.4964 to 4.5180; p=0.3241; Week 52 Δ=2.2791, 95% CI −0.7858 to 5.3441; p=0.1447; Week 65 Δ=2.3716, 95% CI −0.9090 to 5.6521; p=0.1562), and reaching peak at Week 78 (Δ=4.4770, 95% CI 1.0469 to 7.9071; p=0.0106) (**Figure 4C**).

Lower-dose ADMs showed variable, non-significant differences as compared with off-ADM, though these trends fluctuated directionally week to week: Week 26 Δ=2.9570 (95% CI −0.6329 to 6.5468; p=0.1062), Week 39 Δ=2.1267 (95% CI −1.6982 to 5.9516; p=0.2752), Week 52 Δ=1.8053 (95% CI −2.0853 to 5.6959; p=0.3624), Week 65 Δ=3.8286 (95% CI −0.3164 to 7.9736; p=0.0702), Week 78 Δ=−0.1714 (95% CI −4.5115 to 4.1687; p=0.9382).

As with SWR, the higher-dose ADM group in the SDMT measure showed small differences early in the study that became significant at later timepoints as compared with the off-ADM group: Week 26 Δ=0.4490 (95% CI −0.9623 to 1.8603; p=0.5322), Week 39 Δ=1.2399 (95% CI −0.2131 to 2.6929; p=0.0943), Week 52 Δ=0.8626 (95% CI −0.5922 to 2.3174; p=0.2446), Week 65 Δ=1.7828 (95% CI 0.2647 to 3.3008; p=0.0214), Week 78 Δ=2.0564 (95% CI 0.5070 to 3.6059; p=0.0094) (**Figure 4D**). Lower-dose ADMs showed no significant differences as compared with the off-ADM group, with directionally variable effect sizes up to Week 65: Week 26 Δ=−0.1906 (95% CI −1.9787 to 1.5974; p=0.8342), Week 39 Δ=1.2006 (95% CI −0.6354 to 3.0365; p=0.1995), Week 52 Δ=1.1363 (95% CI −0.6994 to 2.9720; p=0.2245), Week 65 Δ=1.7237 (95% CI −0.1880 to 3.6353; p=0.0771). At Week 78, lower dose ADMs resulted in a statistically significant difference as compared with off-ADMs Δ=2.0289 (95% CI 0.0807 to 3.9772; p=0.0413).

For TMS, where more negative Δ indicates less motor worsening off-ADM, higher-dose ADMs showed greater motor worsening as compared with off-ADM, with significant differences emerging from mid-trial: Week 26 Δ=−0.4044 (95% CI −2.3350 to 1.5262; p=0.6808), Week 39 Δ=−1.7885 (95% CI −3.9376 to 0.3607; p=0.1027), Week 52 Δ=−2.9668 (95% CI −5.1804 to −0.7531; p=0.0087), Week 65 Δ=−2.2289 (95% CI −4.5913 to 0.1336; p=0.0644), Week 78 Δ=−2.8656 (95% CI −5.3934 to −0.3377; p=0.0264) (**Figure 4E**). At all visits, lower-dose ADMs showed no significant differences as compared with the off-ADM cohort: Week 26 Δ=−1.0937 (95% CI −3.6181 to 1.4307; p=0.3950), Week 39 Δ=−0.3112 (95% CI −3.0779 to 2.4555; p=0.8252), Week 52 Δ=−1.9773 (95% CI −4.8165 to 0.8619; p=0.1719), Week 65 Δ=−0.7030 (95% CI −3.7227 to 2.3167; p=0.6477), Week 78 Δ=−1.0393 (95% CI −4.2671 to 2.1885; p=0.5274).

Overall, higher-dose ADM exposure was associated with greater worsening over time as compared with off-ADM. In contrast, lower-dose ADM exposure tracked closely with off-ADM across outcomes.

**References:**

[1]	Siafis S, Wu H, Wang D, et al. Antipsychotic dose, dopamine D2 receptor occupancy and extrapyramidal side-effects: a systematic review and dose-response meta-analysis. *Mol Psychiatry* 2023; 28: 3267–3277.

[2]	Torrisi SA, Laudani S, Contarini G, et al. Dopamine, Cognitive Impairments and Second-Generation Antipsychotics: From Mechanistic Advances to More Personalized Treatments. *Pharmaceuticals* 2020; 13: 365.

[3]	Geva M, Goldberg YP, Schuring H, et al. Antidopaminergic Medications and Clinical Changes in Measures of Huntington’s Disease: A Causal Analysis. *Mov Disord*.

[4]	Tan AM, Geva M, Goldberg YP, et al. Antidopaminergic medications in Huntington’s disease. *J Huntingt Dis* 2025; 18796397241304312.

[5]	Bonelli RM, Niederwieser G, Tribl GG, et al. High-dose olanzapine in Huntington’s disease: *Int Clin Psychopharmacol* 2002; 17: 91.
